# Improving respiratory virus forecasts by capturing the effects of school holidays, public holidays and seasonal changes in transmission

**DOI:** 10.64898/2026.11.15.26367964

**Authors:** Ruarai J. Tobin, Oliver Eales, Michael J. Plank, Dylan J. Morris, James M. McCaw, Freya M. Shearer

**Affiliations:** Infectious Disease Dynamics Unit, Melbourne School of Public Health, The University of Melbourne, Australia; School of Mathematics and Statistics, The University of Melbourne, Australia; Infectious Disease Ecology and Modelling, The Kids Research Institute Australia, Australia; School of Mathematics and Statistics, The University of Canterbury, New Zealand; Te Pūnaha Matatini, New Zealand; School of Mathematical Sciences, Adelaide University, Australia

## Abstract

Respiratory viruses impose a substantial burden of morbidity and mortality globally, and models that forecast this burden are widely used to support public health decision-making. However, the public health utility of these forecasts can be limited by poor predictive accuracy. In this work, we show how capturing the effects of school holidays, public holidays and seasonal changes in transmission can improve respiratory virus forecasting performance. We construct a Bayesian renewal model where the effective reproduction number *R*(*t*) is composed of log-additive components including school and public holiday effects and an annually-periodic Gaussian process. The model also allows for the relative rate of reporting to vary with school and public holidays. We use this model to forecast respiratory syncytial virus (RSV), influenza and SARS-CoV-2 case incidence across a 28-day horizon over five jurisdictions in Australia. Applied to the 2025 winter season, our forecasts outperform a random-walk baseline forecast across all settings. We find that the inclusion of holiday and seasonal transmission effects improved forecasting performance across the three pathogens. We show that the school holidays are associated with population-wide reductions in *R*(*t*) on the order of 5–15% but are generally not associated with changes in reporting. Collectively, our results provide support for the inclusion of holiday and seasonal effects in future respiratory virus forecasting models.

## Introduction

The global health burden of respiratory viruses is substantial, with influenza virus and respiratory syncytial virus (RSV) alone estimated to lead to millions of hospital-isations and hundreds of thousands of deaths every year (Iuliano et al. 2018; Savic et al. 2023; Mazur et al. 2024; Hanage and Schaffner 2025). While the future patterns of SARS-CoV-2 transmission remain unclear, the virus continues to spread and cause significant morbidity and mortality (Australian Bureau of Statistics 2026; Zhang et al. 2026; Carstens et al. 2026). Forecasts can provide intelligence to support public health decision-makers as they prepare for and respond to epidemics of respiratory viruses (Muscatello et al. 2017; Johnson et al. 2022; Christen et al. 2026), and a range of forecasting programs have been launched internationally to produce and share such forecasts (Henderson et al. 2026; Gozzi et al. 2026; Hines et al. 2026; MacArthur et al. 2026). Despite substantial advances made in the field over the past decade, improving the performance of respiratory virus forecasts remains an active area of research.

The dynamics of seasonal respiratory virus transmission are the product of complex interactions between patterns in human behaviour, viral transmissibility changes, the build-up and loss of immunity, and demographic turnover (Altizer et al. 2006; Eggo et al. 2016; Bents et al. 2026; Rubin et al. 2026; Tobin et al. 2026). The relationship between these transmission dynamics and observed respiratory virus activity can be further complicated by biases in the observation process (Eales et al. 2024). For case incidence time-series — a common indicator of respiratory virus activity (e.g. Broberg et al. 2018; Hassall et al. 2026), and correspondingly a common target of respiratory virus forecasts (e.g. Song et al. 2016; Bracher et al. 2022; Moss et al. 2026) — the rate at which individuals are tested and infections are detected in case data may vary substantially over time (Moss et al. 2019; Baltrusaitis et al. 2022; Eales et al. 2023). Despite these complexities, patterns in how case incidence data changes over time are still able to be identified, and capturing these patterns within forecasting models provides an opportunity for the improvement of forecast performance. For example, it is regularly observed that, in temperate regions, the incidence of many respiratory viruses is greatest in wintertime and lowest in summertime (Altizer et al. 2006; Moriyama et al. 2020; Kronfeld-Schor et al. 2021). While this pattern is most obvious in temperate regions, similar behaviour can be identified in sub-tropical and tropical regions, particularly for RSV (Lam et al. 2019; Li et al. 2019; Thongpan et al. 2020; Lee et al. 2023). In addition (while not examined in this work), the rate of detection and reporting of respiratory virus infections could also vary seasonally across each year (Moss et al. 2019).

Associations have also long been drawn between the occurrence of school holidays and changes in the reported incidence of respiratory viruses (Stallybrass 1928; Fellowes and Proctor 1973; Storr and Lenney 1989). Past research has primarily attributed these changes to a reduction in the rate of transmission between children (Cauchemez et al. 2008; Eames et al. 2011; Eggo et al. 2016; Ewing et al. 2017; Golding et al. 2023), although changes in healthcare-seeking behaviour during the school holidays could also be involved. The occurrence of public holidays has also been associated with reductions in respiratory virus case incidence, largely due to short-term changes in the rate of testing and reporting (Noah 1989; Lee et al. 2023). However, since public holidays are also associated with changes in the rate of contact between individuals (Van Hoang et al. 2021; Son et al. 2025), their occurrence presumably also has an impact upon transmission.

In this work, we develop a Bayesian renewal model to forecast respiratory virus case incidence. Our model captures the effects of school holidays, public holidays and seasonal changes in transmission upon the effective reproduction number *R*(*t*), as well as the effects of school and public holidays upon the relative rate of case reporting. We apply the model to retrospectively forecast daily RSV, influenza and SARS-CoV-2 case incidence across the 2025 epidemic season in five Australian jurisdictions over a four-week time horizon. We evaluate performance and examine how this performance is affected by the inclusion of different model components. Using the model fit from the final round of forecasts (which utilises the most complete dataset), we examine the estimated transmission and reporting effects, including the effect of school holidays and season on *R*(*t*).

## Results

### The forecasting model

Our forecasting model estimates the effective reproduction number *R*(*t*) by decomposing the quantity log-additively into school and public holiday effects, an annuallyperiodic seasonal Gaussian process, and a non-periodic noise Gaussian process (Figure 1A). We relate this *R*(*t*) to infection incidence through a renewal process (Cori et al. 2013; Abbott et al. 2020), and infection incidence to case incidence via a delay convolution. This case incidence may then further be modified by school and public holiday reporting effects. To produce forecasts, we project *R*(*t*) ahead 28 days, and from this simulate future infection incidence and case incidence. A full description of the model is provided in the Methods.

To illustrate this approach, we present a forecast that was produced across influenza data from Victoria at a time of increasing case incidence, with school holidays occurring during the forecast horizon (Figure 1). In this example, the combined effect of the seasonal Gaussian process and the estimated school holiday transmission effect yields a projected reduction in *R*(*t*) to below one in the immediate horizon of the forecast, and we therefore project a peak in case incidence. The case incidence that was eventually observed falls largely within the central 50% credible intervals predicted. While we estimated no substantial effect of the school holidays upon reporting here, the effect of the King’s Birthday public holiday on the 9th of June is visible (with approximately a halving in the relative rate of reporting).

**Figure 1:**
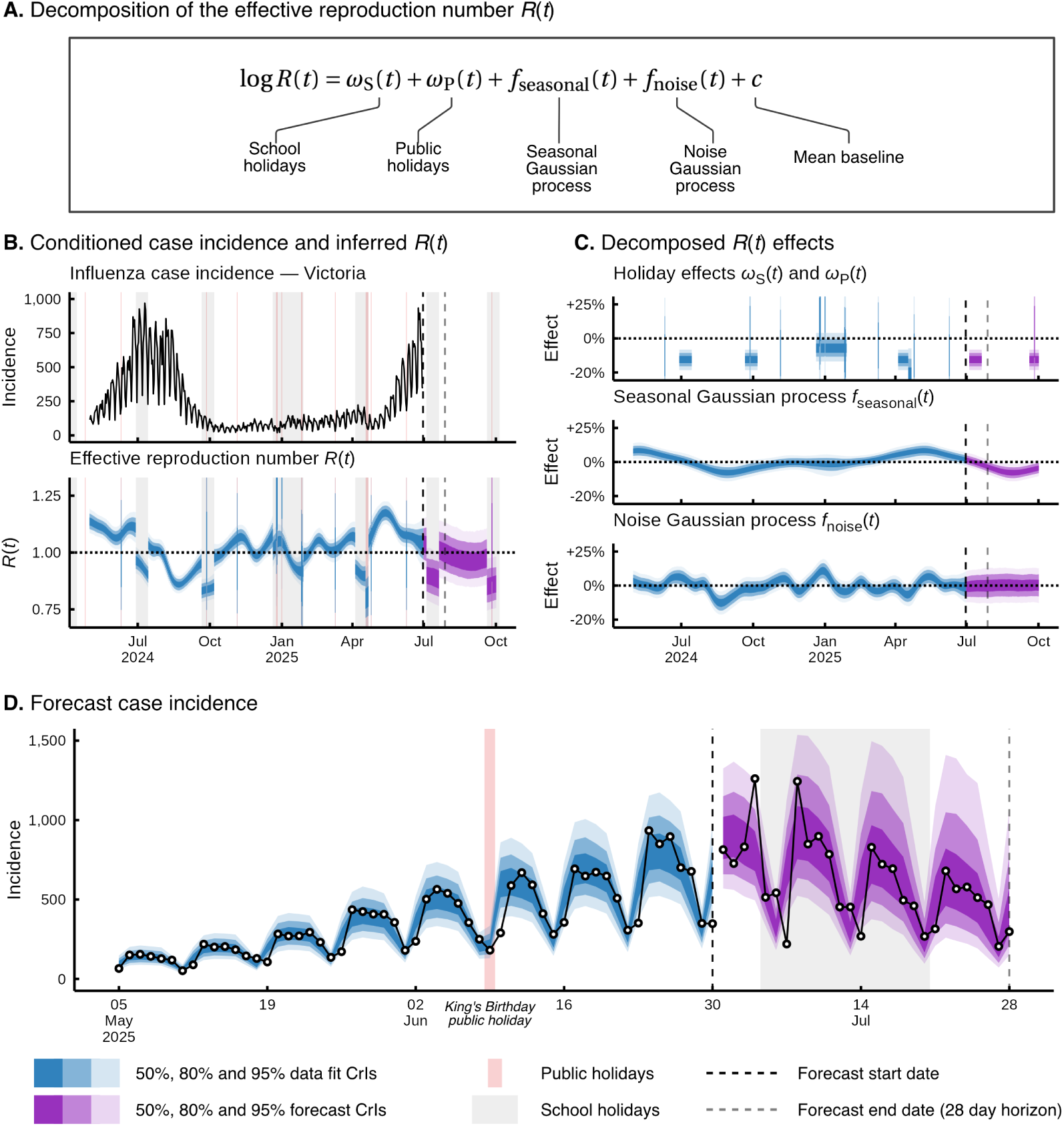
An example forecast produced using our forecasting model (forecast round 2025-07-03). **A**. The *R*(*t*) decomposition model used to capture and forecast case incidence. **B**. Daily influenza case incidence data from Victoria, Australia (note that the total conditioned case incidence includes data back to January 2023, Supplementary Figure 3) and the estimated effective reproduction number *R*(*t*). **C**. The components that make up *R*(*t*), with percentage change calculated as the exponent of the estimated effect minus one. Combined holiday effects are calculated as the product of the school and public holiday effects. **D**. The final forecast case incidence. The case incidence that was eventually observed has been plotted right of the forecast start date. The effects of the King’s Birthday public holiday are visible in a modelled reduction in the relative rate of reporting on that day.

### Forecasting performance

We produced 28-day-ahead forecasts of RSV, influenza and SARS-CoV-2 daily case incidence across five jurisdictions in Australia, with a total of 510 forecasts retrospectively produced across the 2025 epidemic season (approx. February–December). The five jurisdictions were selected to capture a range of epidemiological settings (with typical seasons in each exhibiting distinct transmission dynamics, Supplementary Figures 2–4). Performance was assessed using the continuous ranked probability score (CRPS) across log-transformed counts. Forecasting performance was generally best in the jurisdictions with larger population sizes of New South Wales (NSW, 8.6M residents), Queensland (QLD, 5.7M residents) and Victoria (VIC, 7.1M residents), while performance in the Australian Capital Territory (ACT, 0.5M residents) and the Northern Territory (NT, 0.3M residents) was generally poorer (Figure 2A, Australian Bureau of Statistics 2025). Forecasts of RSV in NSW performed consistently well, with relatively low CRPS across periods of increasing, peak and decreasing epidemic activity (Figure 2B). In contrast, RSV forecasting performance for VIC was limited during increasing and peak activity due to underprediction (forecasts 5–11 in Supplementary Figure 17). Peaks in RSV case incidence coincided with the onset of school holidays in VIC, ACT, QLD and NSW, and forecasting performance tended to be good across these holiday periods (e.g. forecasts 12–15 in Supplementary Figure 13).

**Figure 2:**
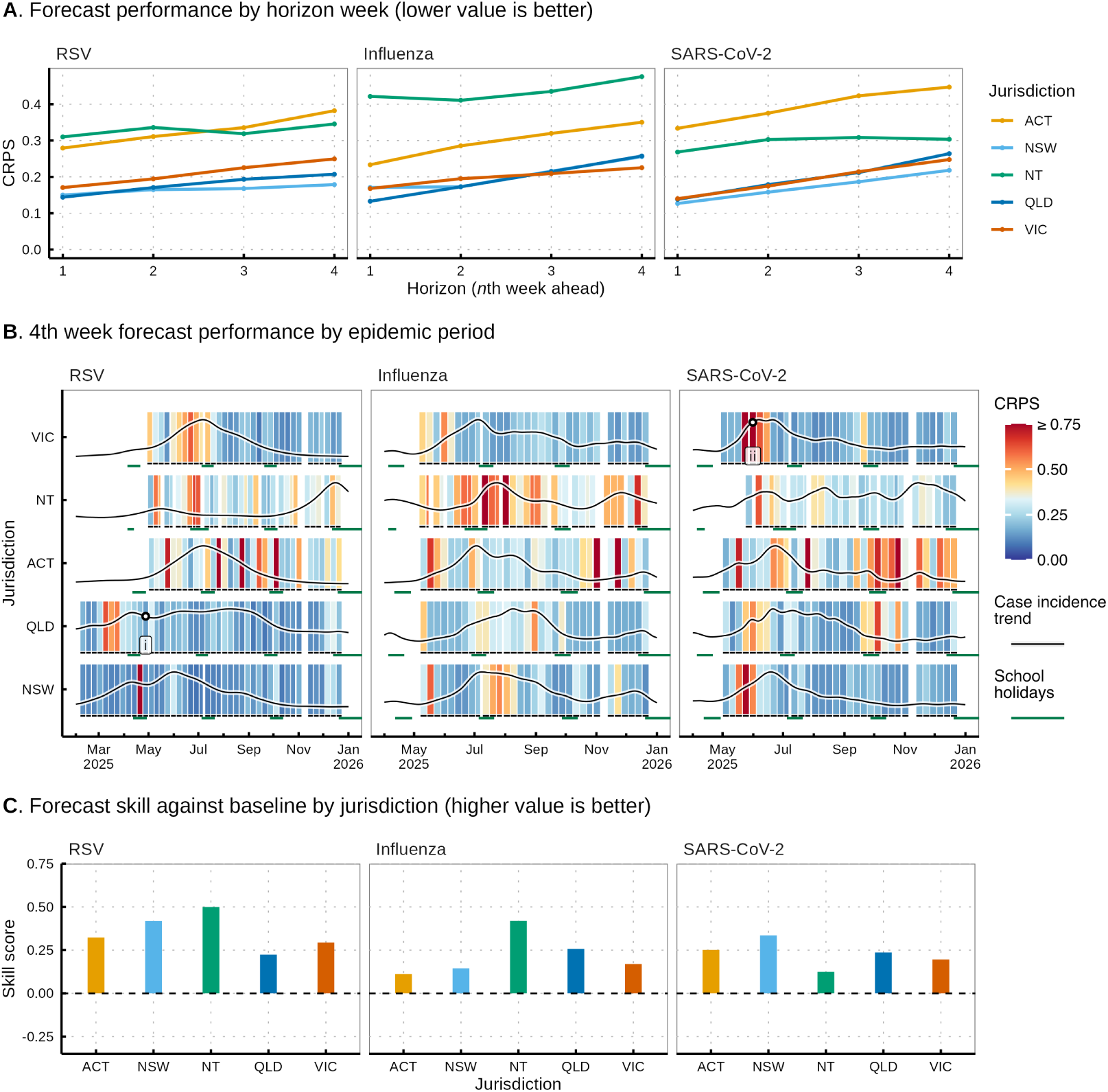
Performance of our forecasting methodology as retrospectively evaluated over the 2025 winter epidemic season in Australia. **A**. The performance of our forecast by horizon week, measured using the continuous ranked probability score (CRPS). A lower value indicates better forecasting performance. **B**. The performance of our forecast at the 4th week horizon (i.e. 22–28 days ahead). STL (Seasonal and Trend decomposition using LOESS) smoothed case incidence is plotted over the CRPS (with scores aligned to the corresponding week of data) to allow for comparison of performance to the underlying epidemiological activity. Where multiple forecasts were produced for the same date (e.g. due to a delay in the receipt of data), the CRPS of the earlier (longer-horizon) forecast is plotted on top. The two labelled forecasts (i, ii) are illustrated in Figure 3. **C**. The relative performance of our forecast as compared to the baseline random-walk model. Skill scores were calculated over the full 28-day-ahead horizon using CRPS over log-counts. A skill score above zero indicates that our forecasts outperform the baseline model, with higher values indicating a greater degree of relative performance.

Forecasting performance for influenza and SARS-CoV-2 was mixed. We found that, similar to RSV, the best overall CRPS was measured for forecasts in the larger jurisdictions (Figure 2A). For influenza, overall performance was often good during the period of epidemic growth early in the season (Figure 2B), but worsened substantially in the weeks following epidemic peaks due to an unexpected late season wave of subclade K influenza A(H3N2) (Dapat et al. 2025), with forecasts tending to predict decreasing case incidence where true incidence remained steady (e.g. forecasts 12–14 in Supplementary Figure 14B). Unlike for RSV, the school holidays were less regularly associated with peaks in epidemic activity for influenza, although peaks may have coincided with school holidays for ACT, NSW and VIC (Supplementary Figure 3). For SARS-CoV-2, transmission dynamics across our study period were dominated by the emergence of the NB.1.8.1 Omicron variant (Uriu et al. 2025), which produced noticeable increases in incidence across all jurisdictions except NT (Supplementary Figure 4). Forecasting performance for SARS-CoV-2 was poor during this period of growth, but improved in the period following peaks in epidemic activity.

Our forecasts outperformed a baseline random-walk forecasting model (see Methods), with overall skill scores identified as being above zero (i.e. better than the baseline) across all pathogen-location pairs (Figure 2C). This skill was best where case incidence was rapidly increasing or decreasing and our forecasts accurately predicted this change (Supplementary Figure 7A). In contrast, skill was worst where case incidence was stable — and thus accurately captured by the baseline model — and our forecasts incorrectly predicted growth or decline in cases (e.g. across the subclade K influenza epidemic as described above). Forecast skill was generally good in jurisdictions with lower population sizes, indicating that the relatively poorer performance as measured by CRPS in these settings may be reflective of the inherent difficulty in forecasting at lower case counts.

Where our model failed to capture underlying epidemiological dynamics — such as in the early growth phase of the SARS-CoV-2 wave in Victoria — worsened performance tended to be reflected across all four horizon weeks (Supplementary Figure 5). In contrast, a singular week of data that was poorly predicted by four subsequent forecasts more often reflects an unexpected data anomaly (e.g. RSV forecasts 11–14 in Supplementary Figure 14A, where case incidence was reported as zero across the Easter holiday period).

### Effect of the holiday and seasonality model components upon performance

To examine how different components of the model contributed to the forecasting performance, we illustrate two example forecasts and the components underlying their projected *R*(*t*) (Figure 3). First, we examine a forecast of RSV in Queensland, this being a typical example of a well performing forecast (Figure 3, panel **i**). This forecast was produced at a time of gradually increasing case incidence and immediately prior to the autumn school holidays. We see that the predicted case incidence accurately captured what was eventually observed, and uncertainty in the forecast increased only marginally across the four week horizon. Examining the model components underlying *R*(*t*), we see that this accuracy can be attributed to the combined effect of two components. First, the seasonal Gaussian process *f*_seasonal_(*t*) gradually slowed transmission across the forecast horizon, reflecting reductions in transmission that had been observed at the same time in prior years (Supplementary Figure 2). Second, the holiday effects reduced *R*(*t*) even further, again reflecting reductions in transmission that had been observed across the school holidays and Easter in prior years. In addition, the model accurately predicted that a step-change in case incidence reporting would occur across Easter. The same forecast produced without holiday or seasonality effects largely fails to predict a slowdown in transmission across the horizon (Supplementary Figure 6A).

**Figure 3:**
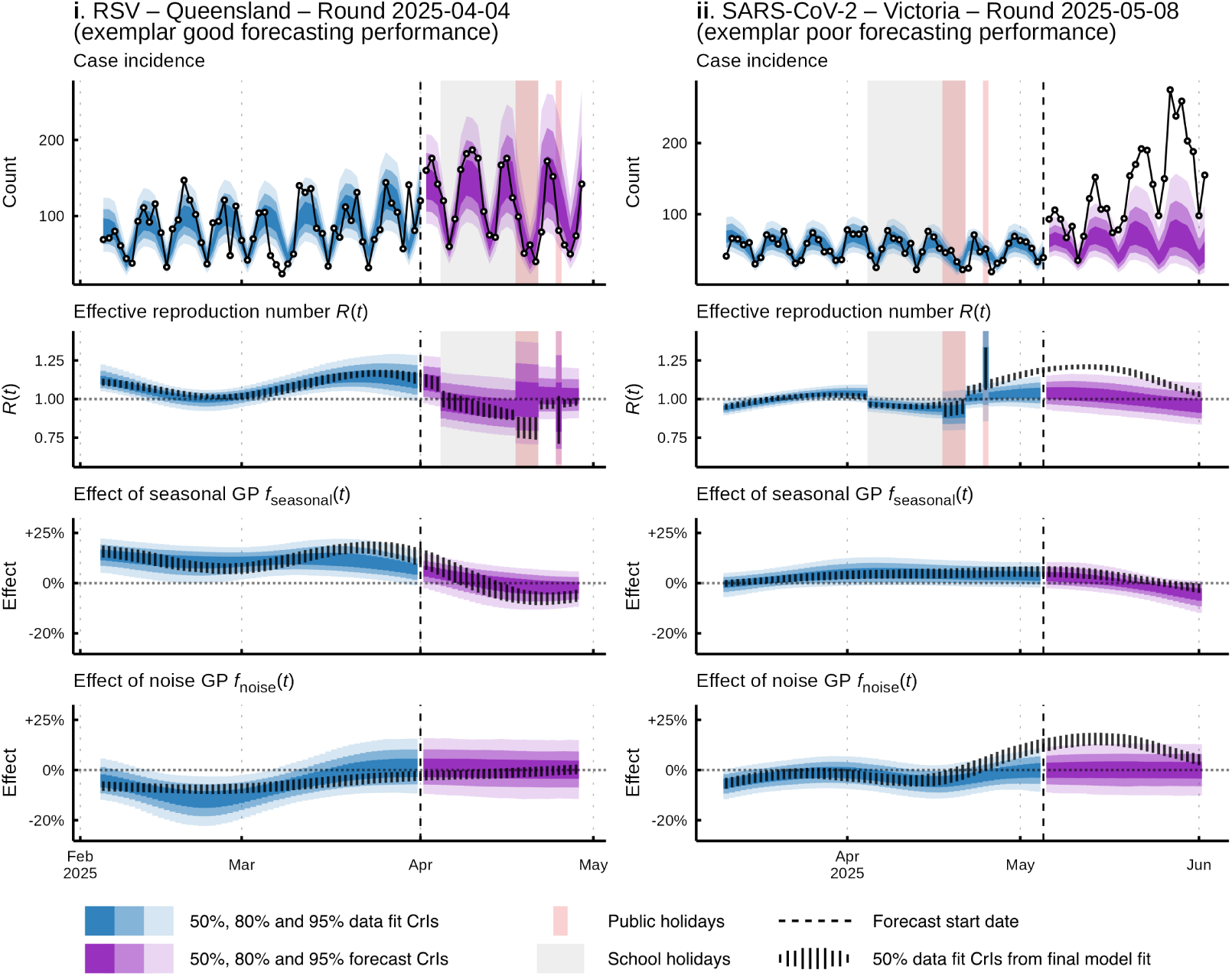
The two example forecasts that are highlighted in Figure 2B (with labels **i** and **ii**), exhibiting good (left, panel **i**) and poor (right, panel **ii**) forecasting performance. Vertical lines indicate the 50% credible intervals taken from the model fit to data from the final forecasting round (round 2025-11-27). Percentage changes due to the seasonal and noise Gaussian process effects were calculated as the exponent of the estimated effect minus one.

In contrast, our forecast for SARS-CoV-2 in Victoria (Figure 3, panel **ii**) substantially underpredicted the case incidence that was eventually observed due to an unexpected increase in case incidence. At the time of forecasting (2025-05-08), there was no signal in the case incidence data that *R*(*t*) would increase, with the seasonal Gaussian process instead predicting that *R*(*t*) would slightly decrease as in prior years. Instead, a substantial increase in transmission occurred across the forecasting horizon due to the emergence of the NB.1.8.1 Omicron variant (Uriu et al. 2025). The model fit produced for the final forecast round (i.e. conditioned on the entire time-series of data including the NB.1.8.1 epidemic) later attributed this increase in *R*(*t*) to a sustained increase in the noise Gaussian process *f*_noise_(*t*) above what was originally predicted.

In a sensitivity analysis, we found that overall forecasting performance was best where both holiday effects (including the effects of school and public holidays on *R*(*t*) and reporting) and the seasonal Gaussian process were included in the model for RSV and influenza (Figure 4). In contrast, performance for SARS-CoV-2 was largely unaffected by the inclusion of the seasonal Gaussian process. This difference is reflective of the epidemiological dynamics of SARS-CoV-2 observed across 2023– 2025, where a consistent seasonal pattern has yet to emerge (Supplementary Figure 4).

**Figure 4:**
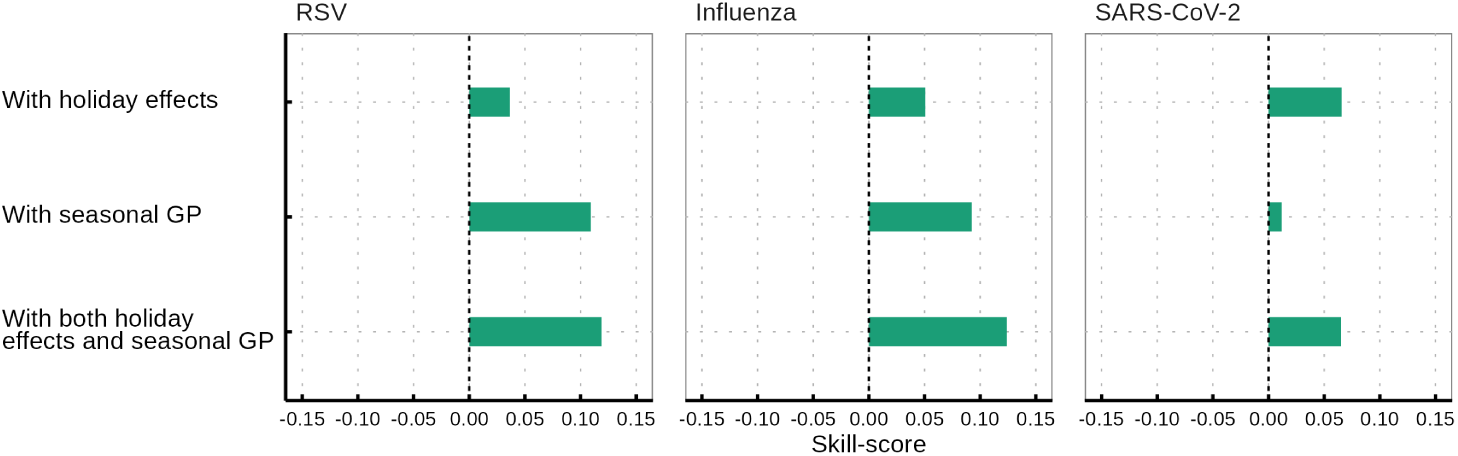
The relative 4th week forecasting performance of the model in comparison to a null model with neither holiday effects (including school and public holiday effects on ascertainment and *R*(*t*)) or the seasonal Gaussian process.

The performance impact of these model components was dependent upon the epidemiological context (Supplementary Figure 7B). For example, as the seasonal Gaussian process contributed to incorrect predictions of epidemic decline during the subclade K epidemic wave (as discussed above), the model that included neither the holiday effects or the seasonal Gaussian process performed best across this period. However, around peaks in influenza epidemic activity, forecasting performance was often supported by the inclusion of these components. For example, the model including both holiday effects and the seasonal Gaussian process confidently (and correctly) predicted a peak in influenza case incidence in Victoria, whereas the model without these components suggested that incidence could continue to increase (Supplementary Figure 6B).

For influenza and RSV, we found that the majority of the performance benefits of including school and public holiday effects in the model could be attributed to their effect on transmission (as opposed to their effect on reporting). Removal of the holiday transmission effects (across both school and public holidays) degraded forecasting performance more than removing the holiday reporting effects for these two pathogens (Supplementary Figure 8), while for SARS-CoV-2, removing either effect produced a similar reduction in performance. Further, the performance benefits of including holiday effects were primarily attributable to the school holiday components for influenza and SARS-CoV-2 (with the inclusion of the school holiday effects marginally reducing performance for RSV). In contrast, the performance benefits of the holiday effects were attributable largely to the public holiday components for RSV. We found that removal of the delay between infection and reporting marginally improved performance for influenza (with an approximately 1% reduction in CRPS), while performance for SARS-CoV-2 and RSV were effectively unchanged (Supplementary Figure 8). The potential benefits of including this delay may require that estimates of the delay distribution can be provided ahead of time, rather than inferred from case incidence data alone as we have done here.

### Estimated school and public holiday transmission and reporting effects

We find that the between-term school holidays were associated with a substantial reduction in transmission (Figure 5A). These estimates were taken from the final forecasting round (round 2025-11-27, with this round having the most complete set of data considered in the study). Across the five jurisdictions and three pathogens, the between-term school holiday effects on *R*(*t*) were identified to be on the order of -5 to -15%, with 95% credible intervals falling entirely below zero for a number of jurisdiction-pathogen pairs (Supplementary Table 1). These transmission effects were most apparent for influenza and in the jurisdictions with larger population sizes. In contrast, the effects of the summer holidays on transmission were substantially less certain with no clear trend evident between different jurisdictions (Supplementary Figure 9A). The effect of the between-term school holidays on case reporting was less certain, with 95% credible intervals only falling below zero for influenza and SARS-CoV-2 in NSW. Similarly, the effect of summer school holidays on reporting was highly uncertain with only minimal evidence of a reduced rate of reporting (Supplementary Figure 9B).

**Figure 5:**
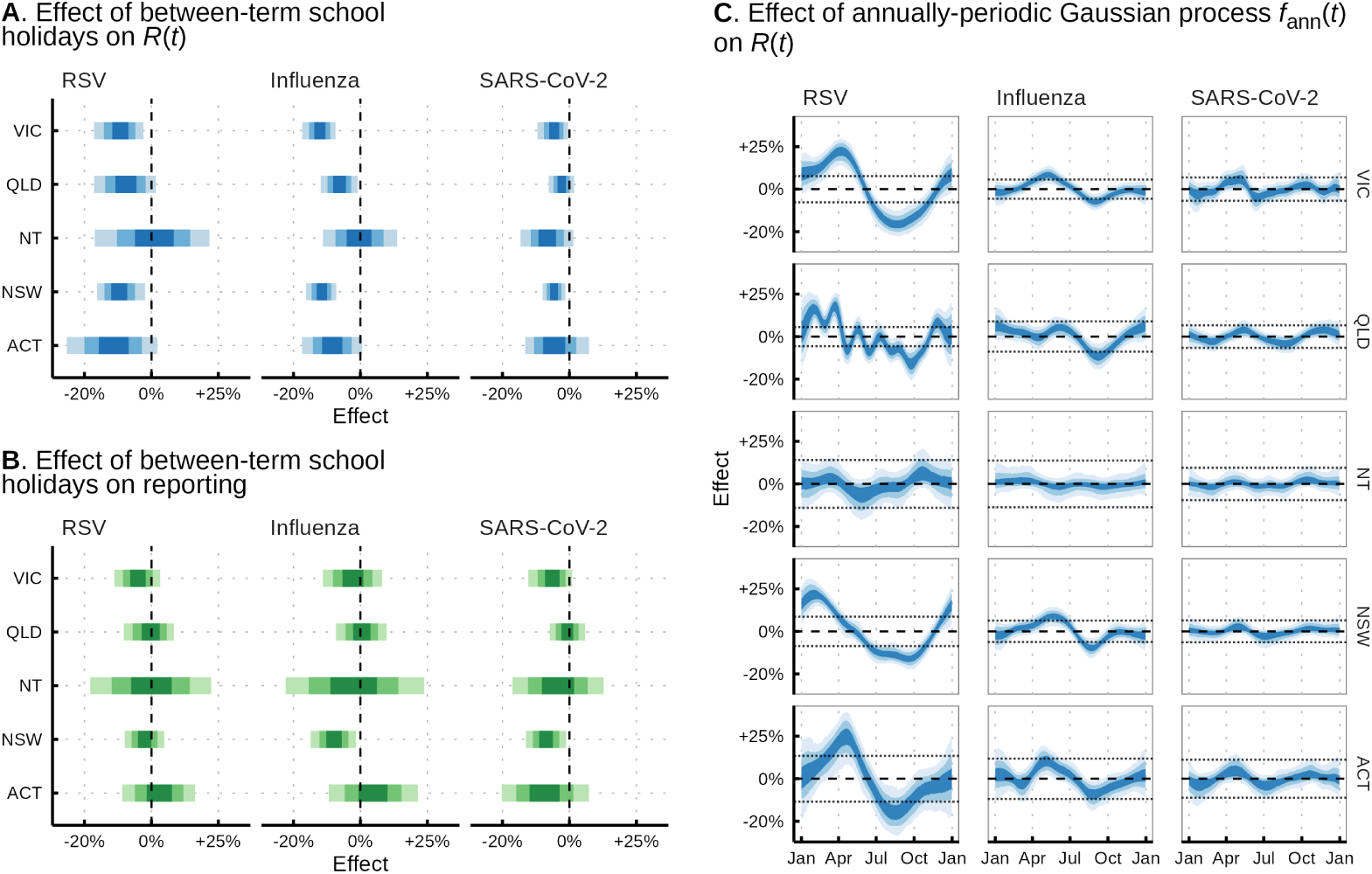
Estimated between-term school holiday effects and seasonal Gaussian process terms for each jurisdiction/pathogen considered in the study, taken from the final model fit (forecast round 2025-11-27). For each, the 50%, 80% and 95% credible intervals are displayed. Percentage change effects were calculated as the exponent of the estimated effect minus one. **A**. The effect of between-term school holidays on *R*(*t*). **B**. The effect of between-term school holidays on the relative rate of reporting. **C**. The effect of the seasonal Gaussian process on *R*(*t*) across one year. The dotted horizontal lines indicate the typical amplitude (standard deviation) of the noise Gaussian process *f*_noise_.

The effects of public holidays were found to behave in an opposite manner to that of school holidays, with their influence on *R*(*t*) uncertain but their influence on the relative rate of reporting well identified and of substantial magnitude (Supplementary Figure 10). Only a few public holidays were found to have an effect on *R*(*t*) with a 95% credible interval that did not overlap zero, and these may have occurred by chance alone given the large number of combinations of public holidays, jurisdictions and pathogens tested (Supplementary Figure 10A). In contrast, the effects of public holidays on case reporting were found to consistently lead to a reduction in the relative rate of reporting (Supplementary Figure 10B). In addition, similarities in the effect of certain public holidays were identified between different settings (e.g. the King’s Birthday holiday was consistently associated with a reduction in reporting of around 20–50%).

### Estimated seasonal transmission effects

The effect of the seasonal Gaussian process *f*_seasonal_(*t*) on *R*(*t*) was of the largest magnitude for RSV, where we observed annual increases and decreases of around 20–25% for all jurisdictions except the Northern Territory (Figure 5C). The effect grew earliest in the year for the jurisdictions with sub-tropical population centres of New South Wales and Queensland, while increases began later in the year for the temperate jurisdictions of Victoria and the Australian Capital Territory. The effects of *f*_seasonal_(*t*) for influenza were of lower amplitude, with increases and decreases on the order of 5–10%, which is comparable to the amplitude of the noise Gaussian process term *f*_noise_(*t*) in all jurisdictions. The effect of *f*_seasonal_(*t*) was found to be the least substantive for SARS-CoV-2, although a small increase in *R*(*t*) of around 5% around April was consistently identified across jurisdictions.

## Discussion

We have presented a Bayesian respiratory virus forecasting model where we capture recurring variation in the effective reproduction number *R*(*t*) through school and public holiday effects and an annually-periodic seasonal Gaussian process. Our model further captures recurring changes in the relative rate of reporting that may arise across school and public holidays. We have shown that our model exhibits excellent forecasting performance in contexts where these effects are predictive of epidemic dynamics. This predictive capability was most evident in forecasts of RSV and in the jurisdictions within Australia with larger population sizes. Even for pathogens where our model was not able to capture the primary drivers of epidemic growth (e.g. the emergence of new antigenic variants of influenza or SARS-CoV-2), performance was best overall where both holiday and seasonal effects were included in the model. We also estimated the effects of holidays on respiratory virus dynamics and found the strongest effects to be those of the between-term school holidays on *R*(*t*) and of the public holidays on the relative rate of reporting.

Across the five jurisdictions in Australia and the three respiratory viruses we considered in this work, we found that the school holidays were often associated with changes in transmission but less so with changes in reporting. The transmission effects that we estimated — on the order of a 5–15% reduction in *R*(*t*) — are consistent with prior work. For example, Ewing et al. (2017) estimated that the *R*(*t*) of influenza-like-illness reduced by approximately 15% across the winter holidays in the US, while Wang et al. (2026) estimated a 14% reduction in the *R*(*t*) of influenza A(H1N1) across the 2009 summer holidays in China. Our population-level estimates are naturally lower than age-specific estimates of reductions in transmission between children during the school holidays, which have been reported to be on the order of 25–45% (Cauchemez et al. 2008; Eggo et al. 2016). Our estimates are valuable in informing our understanding of respiratory virus dynamics, as even small recurring changes in transmission can have an outsized effect on quantities such as final size, peak size and peak timing (Altizer et al. 2006; Eames 2014; Tobin et al. 2026).

Our finding that public holidays have substantial effects on reporting aligns with prior research internationally that has found substantial changes in the rate of testing and reporting across public holidays (Lee et al. 2023; Rumack 2023). While it has been previously reported that public holidays can lead to reductions in contact rates between individuals (Van Hoang et al. 2021; Son et al. 2025), we did not find convincing evidence of any corresponding change in the rate of transmission. However, such changes may be difficult to infer from case incidence data alone, as delays between infection and reporting are likely to obscure the effect of a single-day change in transmission. Incorporation of public holiday effects in operational forecasting models may be better achieved through use of additional data streams such as ongoing, frequent contact surveys (Golding et al. 2023).

While the inclusion of all holiday effects in our model improved forecasting performance, we found that the impact was more mixed when broken down by type of holiday or type of effect. For example, the inclusion of the school holiday transmission and reporting effects resulted in improved forecasting performance for SARS-CoV-2 and influenza but not RSV, where performance was marginally worse. This difference may be a consequence of RSV cases most often occurring in children younger than school-age (Mazur et al. 2024), so the school holidays presumably impact a smaller fraction of potential transmission events.

The seasonal Gaussian process component of the forecasting model proved most beneficial for forecasting performance in epidemiological contexts where changes in transmission were highly consistent year-on-year. We found the inclusion of this component had the greatest performance benefit for RSV, followed by influenza, with only marginal benefit found for SARS-CoV-2. The patterns of RSV transmission are highly seasonal, with the timing of epidemic growth, peak and decline relatively consistent from one year to the next. These stable dynamics were very well captured by our periodic Gaussian process, enabling accurate prediction of RSV case incidence.

While influenza epidemics typically recur at similar times year-to-year, the emergence of new viral strains can produce dynamics that are not well captured by the periodic Gaussian process. This was evident in our application to the 2025 influenza season in Australia, where the unexpected emergence of the influenza A(H3N2) subclade K (Dapat et al. 2025) produced a prolonged period of heightened influenza case incidence. As our model does not capture strain or sub-type-level variation in growth (Davies et al. 2021; Eales et al. 2025), the model instead incorrectly predicted that case incidence would decline following the initial peak. However, the emergence of subclade K was an unusual epidemiological event, and it is possible that the performance of our model in forecasting influenza would be greater in a more typical season.

This non-periodicity was most evident for SARS-CoV-2, where epidemics have not followed a stable seasonal pattern across the 2023–2025 years. While these nonperiodic dynamics limited the model’s ability to predict changes in growth rate ahead of time, our forecasts still proved capable of adapting to these changes once they were evident in the data. Further, the inclusion of the periodic term is not necessarily harmful for overall performance in such non-periodic settings, as the magnitude of the term can automatically be shrunk towards zero during inference if periodicity is able to be ruled out.

Our estimates of the seasonal Gaussian process *f*_seasonal_ should not be interpreted as equivalent to a seasonal forcing term on transmission. Instead, this term likely captures a combined effect of changes in both the rate at which the virus can be transmitted (i.e. seasonal forcing, due to changes in human behaviour or viral transmissibility) and changes in population immunity (with reductions in transmission due to susceptible depletion and increases due to waning immunity). To separate out these components, our model would need to be able to explicitly capture the effects of accumulating immunity (due to exposure) and depleting immunity (due to waning) on the population-scale transmission of respiratory viruses. This remains a difficult task due to the complex dynamics of respiratory virus immunity and our poor understanding of the proportion of infections that are detected in case data.

Accurate forecasts of seasonal respiratory virus burden can aid public health decision-makers in critical areas such as medical workforce scheduling, testing and supply chain planning and public communications (Muscatello et al. 2017; Moss et al. 2018; Frounfelker et al. 2025; Christen et al. 2026). A large number of respiratory virus forecasting programs are currently running (e.g. Gozzi et al. 2026; Hines et al. 2026; Henderson et al. 2026), and improving the performance of these forecasts is an active field of research. Our results show that capturing the effects of school holidays and seasonal changes in transmission can substantially improve the performance of respiratory virus forecasts. Such improvements may be of the greatest utility around peaks in epidemic activity, which in many settings (including ours), often coincide with the onset of school holidays (Cauchemez et al. 2008; Garza et al. 2013). Our model is currently being used to produce real-time forecasts of RSV, influenza and SARS-CoV-2 across eight states and territories in Australia and in New Zealand on a weekly basis as part of the Australia-Aotearoa Winter Situational Assessment Program, with evaluation to follow. Our results are likely to generalise to respiratory virus forecasting work in other settings, and the inclusion of holiday and seasonal effects should be considered where similar patterns in transmission and reporting can be identified.

## Methods

### Bayesian generative model

We consider the log-transformed effective reproduction number log *R*(*t*) to be made up of five additive components:

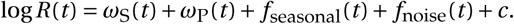

The functions *ω*_S_(*t*) and *ω*_P_(*t*) capture changes in transmission due to the school holidays and public holidays respectively (e.g. due to changes in contact behaviour), while *f*_seasonal_(*t*) captures annually-periodic changes in the transmission rate and *f*_noise_(*t*) captures any variation not explained by the other three terms. The baseline *c* is included to capture any net difference from log *R*(*t*) *=* 0 across the fitting period.

For the school holiday term *ω*_S_(*t*), we estimate two separate effects. The value *ω*_between-term_ captures any changes to transmission across the shorter between-term school holidays, which are two weeks long and occur every (Southern Hemisphere) autumn, winter and spring, while *ω*_summer_ captures transmission changes across the summer holidays, which are approximately six weeks long (and coincide with the Christmas and New Year holiday period in Australia). These effects apply whenever time *t* is in the respective set of time-steps *H*_between-term_ or *H*_summer_. Otherwise, *ω*_S_(*t*) is zero:

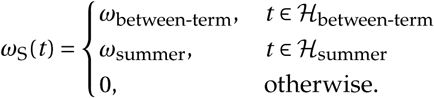

Priors for these parameters (and others) are listed in Table 1.

**Table 1:**
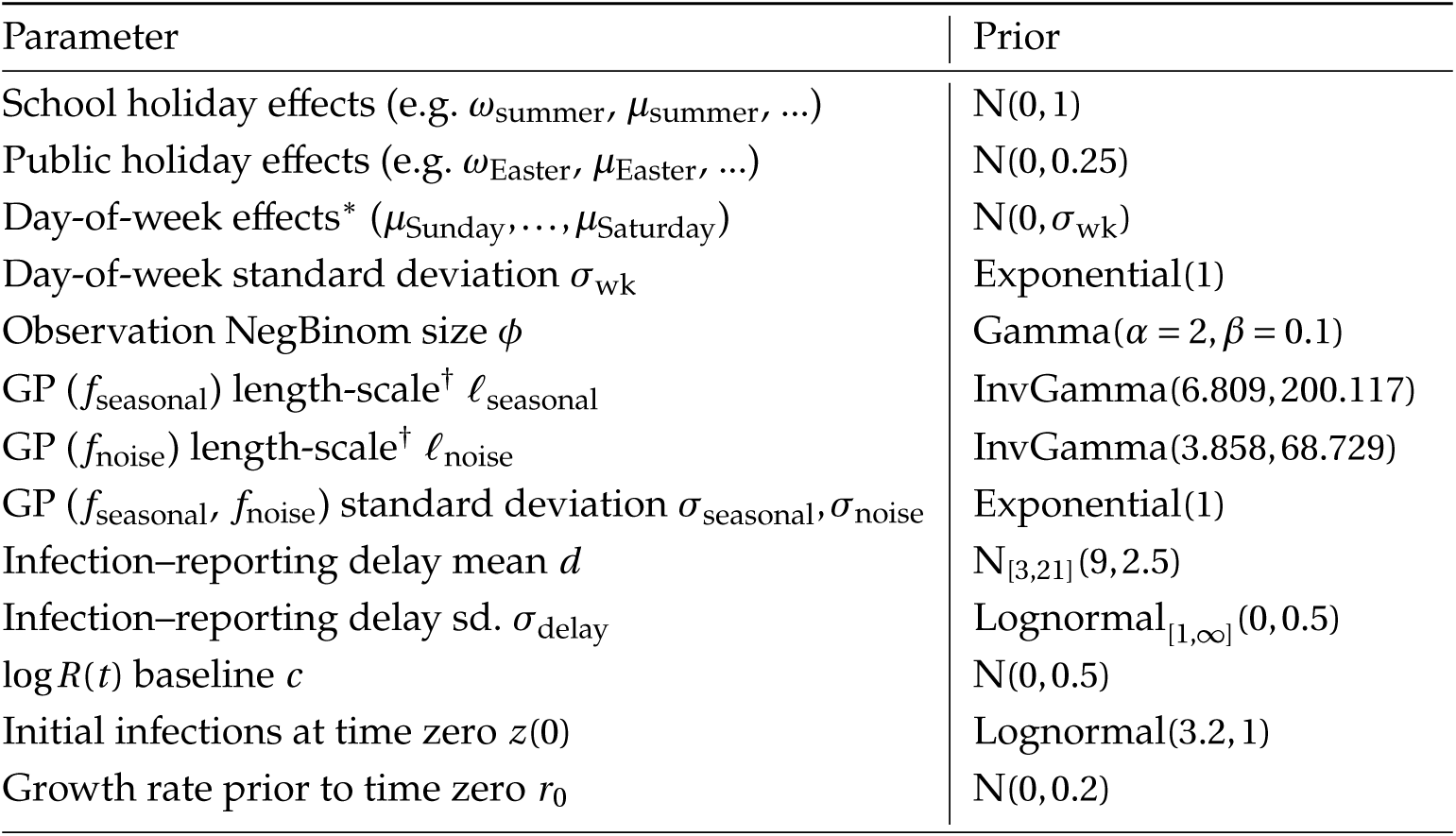
Priors for the Bayesian model. Truncated distributions are indicated with subscript intervals. A *µ* of 3.2 for *z*(0) approximately corresponds to an initial case incidence of 25. Normal distributions are parametrised using mean and standard deviation, and the inverse-gamma distributions are parametrised in terms of shape and rate. *^∗^*: Note that the sum-to-zero constraint applied to the day-of-week effect means these priors are not independent. ^†^: Values specified such that the 99% intervals were approximately [14, 90] days and [7, 90] days respectively as described in the text.

| Parameter | Prior |
|---|---|
| School holiday effects (e.g. ω<sub>summer</sub>, μ<sub>summer</sub>, ...) | N(0, 1) |
| Public holiday effects (e.g. ω<sub>Easter</sub>, μ<sub>Easter</sub>, ...) | N(0, 0.25) |
| Day-of-week effects<sup>*</sup> (μ<sub>Sunday</sub>, …, μ<sub>Saturday</sub>) | N(0, σ<sub>wk</sub>) |
| Day-of-week standard deviation σ<sub>wk</sub> | Exponential(1) |
| Observation NegBinom size ϕ | Gamma(α = 2, β = 0.1) |
| GP (f<sub>seasonal</sub>) length-scale<sup>†</sup> ℓ<sub>seasonal</sub> | InvGamma(6.809, 200.117) |
| GP (f<sub>noise</sub>) length-scale<sup>†</sup> ℓ<sub>noise</sub> | InvGamma(3.858, 68.729) |
| GP (f<sub>seasonal</sub>, f<sub>noise</sub>) standard deviation σ<sub>seasonal</sub>, σ<sub>noise</sub> | Exponential(1) |
| Infection–reporting delay mean d | N<sub>[3,21]</sub>(9, 2.5) |
| Infection–reporting delay sd. σ<sub>delay</sub> | Lognormal<sub>[1,∞]</sub>(0, 0.5) |
| log R(t) baseline c | N(0, 0.5) |
| Initial infections at time zero z(0) | Lognormal(3.2, 1) |
| Growth rate prior to time zero r<sub>0</sub> | N(0, 0.2) |

We estimate a separate effect of each named public holiday in a similar manner. In the case where a public holiday fell on a weekend and was officially observed on the preceding Friday or following Monday, we assumed this could have a different effect on transmission and as such estimated a separate effect here (for example, we estimate one effect for ‘New Year’s Day’ and one for ‘New Year’s Day (Observed)’). Lists of official public holiday and school holiday dates were compiled from online sources with archived copies of reference material made available in the supplementary data. Differences in holiday dates between jurisdictions for 2025 are presented in Supplementary Figure 1. For simplicity of interpretation, the four public holidays defining the Easter period are treated as a single holiday.

The term *f*_seasonal_(*t*) captures any seasonal variation in *R*(*t*) that recurs on a periodic, annual basis. We capture this behaviour by modelling *f*_seasonal_(*t*) as a Gaussian process with a periodic squared-exponential kernel:

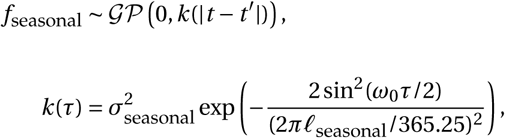

with *ω*_0_ *=* 2*π*/365.25 such that the period of the process is 365.25 days.

Finally, the term *f*_noise_(*t*) captures any variation not more readily explained by the previous three terms. We model this term as a Gaussian process with a Matérn 5/2 kernel:

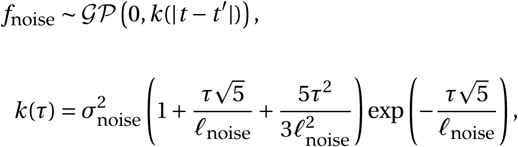

with this kernel selected to capture an intermediate level of smoothness in the noise process.

The latent infection incidence *z*(*t*) is defined as a renewal process (as in Cori et al. 2013; Abbott et al. 2020) across the effective reproduction number *R*(*t*):

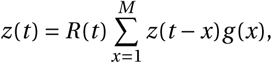

where *g* (*x*) is the probability of an onwards infection occurring after *x* days, i.e. the generation interval. Discretised generation interval distributions were taken to be fixed and specified from prior studies for each of the three pathogens modelled (Cauchemez et al. 2009; Vink et al. 2014; Abbott et al. 2022). The value of *M* was taken to be fixed at 21 days.

To calculate the latent infection incidence as defined above, we must also define *z*(*t*) for the *M* days prior to the first day of data. We do this by assuming that exponential growth/decay in *z*(*t*) occurs across this period:

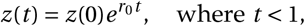

with the initial infection incidence *z*(0) and growth rate *r*_0_ treated as free model parameters (Table 1).

We assume that the mean case incidence *y* (*t*) is a delayed convolution of the latent infection incidence *z*(*t*). The convolution *p*_delay_ captures the delay from the onset of infection to case notification date. This distribution is captured as a Log-normal distribution *X* with a mean of *d* and a standard-deviation of *σ*_delay_. This continuous distribution is truncated at *M* days and discretised across each day 0 through *M −* 1:

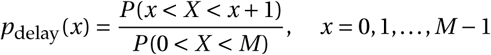

The mean and standard deviation of the untruncated distribution are treated as parameters to be inferred during the fitting process. As the exact timing of infection incidence and thus this distribution is difficult to estimate from case incidence data alone (with only limited information provided by e.g. the timing of public holidays and any corresponding impact on transmission), constrained priors were applied to prevent unrealistically short (or long) delay distributions being inferred (Table 1). Estimates for *p*_delay_ from the final round of forecasts (round 2025-11-27) are presented in Supplementary Figure 11.

In addition, we assume that mean case incidence may be increased or reduced during the school holidays via *µ*_S_(*t*), public holidays via *µ*_P_(*t*), and according to day-of-week via *µ*_wk_(*t*):

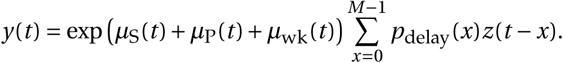

The school and public holiday reporting effects are estimated in the same manner as that taken for their approach on transmission, with separate effects estimated for the summer and between-term holidays and one effect for each named public holiday. For the day-of-week effect *µ*_wk_(*t*), we estimate seven values, one for each day of the week:

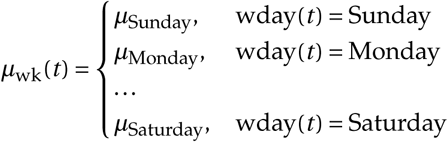

under the sum-to-zero constraint that *µ*_Sunday_ *+µ*_Monday_ *+* . . . *+µ*_Saturday_ *=* 0.

From the mean case incidence *y* (*t*), we assume observed case incidence *Y* (*t*) is drawn from a negative binomial distribution:

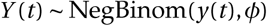

with dispersion (size) parameter *φ*.

The full list of model priors is provided in Table 1. For the Gaussian process length-scale priors, we use the approach suggested in (Betancourt 2017), where an inverse Gamma distribution prior is used with parameters selected such that the 99% interval of the distribution covers a desired range. For the seasonal periodic Gaussian process *f*_seasonal_, the length-scale *ℓ*_seasonal_ is specified such that the 99% interval covers the range [14, 90] (i.e. *P* (14 *ℓ*_seasonal_ 90) 0.99). The probability density of these inverse Gamma priors drops rapidly outside of these regions, in effect preventing the inference of particularly low length-scales (i.e. over-fitting) without imposing a hard cut-off. Similarly, for *f*_noise_, the length-scale *ℓ*_noise_ is specified such that the 99% interval covers the range [7, 90] (with a lower cut-off chosen here for *ℓ*_noise_ under the assumption that the noise process may be more flexible than seasonal changes in transmission).

### Data

Forecasts were produced retrospectively across 33 rounds of historical data from the Australia-Aotearoa Forecasting Hub. These rounds spanned the 2025 winter epidemic season in Australia, with each round consisting of a dataset of case incidence data for RSV, influenza virus and SARS-CoV-2. Use of case data for this study was approved by the Health Sciences Human Ethics Sub-Committee, Office for Research Ethics and Integrity, The University of Melbourne (Application ID 2024-26949-50575-3, “Real-time epidemic analysis and forecasting for decision support”).

Linelisted case data was extracted from the Public Health Event Surveillance System (PHESS) for Victoria and from the National Notifiable Disease Surveillance System (NNDSS) for all other jurisdictions. For Victoria, non-electronic case records were excluded to reduce the impact of reporting delays. From these linelists, aggregate case counts were produced for each forecasting round, with truncation performed as necessary to reduce the potential effect of reporting delays. To reduce the impacts of pandemic-related changes in epidemiological dynamics and reporting on our estimates, daily case incidence before 2023-01-01 was removed for RSV and influenza, and before 2023-03-01 for SARS-CoV-2. This later date for SARS-CoV-2 was chosen due to substantial changes in the availability of PCR testing around the start of 2023 (Department of Health, Australia 2022). This daily case incidence is illustrated in Supplementary Figures 2–4.

In addition to the use of historically produced data for the rounds spanning from May 2025 through to December 2025, additional synthetic rounds were produced by truncating case incidence data. A total of 12 rounds of RSV data were produced for Queensland and New South Wales starting from February 2025 to capture the early increases in case incidence observed in these jurisdictions (see Supplementary Figure 2) and four rounds of RSV, influenza and SARS-CoV-2 data for Victoria in April and May 2025 to capture the period of early epidemic growth and align coverage with other jurisdictions. We produced results across the jurisdictions of the Australian Capital Territory, New South Wales, the Northern Territory, Queensland and Victoria. While case data was available for South Australia, Tasmania and Western Australia, model behaviour was similar to that observed in the Australian Capital Territory and Victoria. As such, we excluded these jurisdictions from the analysis for simplicity of presentation.

### Inference

Model inference was performed in Stan using the No-U-Turn-Sampler (NUTS) with default parameters (Stan Development Team 2026). For the final model fit (where effect estimates were presented, such as in Figure 5), 5, 000 warm-up iterations followed by 20, 000 sampling iterations were performed across four chains to ensure convergence (with *R̂* 1.01 and effective sample sizes over 400 across all parameters). To avoid the computational burden of prolonged sampling, prior rounds were initially fit with 2, 000 warm-up and 2, 000 sampling iterations. If *R̂* was observed to be greater than 1.05 for these fits, fitting was performed with 5, 000 warm-up iterations and the total number of sampling iterations was increased in increments of 20, 000 until *R̂* 1.05 was achieved (although further sampling produced no discernable difference in forecasting performance). Difficult parameter identification was primarily induced by multimodality in the delay distribution parameters and the public holiday effects. The cmdstanr interface was used to perform all sampling (Gabry et al. 2025).

To improve the computational efficiency of model fitting given the long timeseries used in our model (multiple years of data), the Matérn and periodic Gaussian processes were approximated via Hilbert space Gaussian processes (HSGP) and linear low-rank approximations respectively following the approach presented in Riutort-Mayol et al. (2023). The two methods approximate a Gaussian process with a linear combination of *K* basis functions, reducing the computational complexity of inference with Gaussian process priors from *O* (*n*^3^) to *O* (*nK K*), where *n* is the number of time-points evaluated and *K* is typically less than *n*.

Hyper-parameters for the Gaussian process approximations were specified following the guidance provided in Riutort-Mayol et al. (2023). The number of basis functions *K* for the seasonal Gaussian process *f*_seasonal_(*t*) was taken to be fixed at 40. For the noise Gaussian process *f*_noise_(*t*), we calculated the number of basis functions for each model fit using Equation 21 from Riutort-Mayol et al. (2023), with *K* 2.65(*C S*)/7, where *S* is half the number of days in the simulation and we assumed a lower bound on the length-scale of 7 days. Finally, the boundary factor *C* was taken to be 1.25. These hyperparameters were validated to ensure that a highly accurate approximation was produced across the range of length-scales specified by the two priors (Supplementary Figure 12). Stan implementations defining the HSGP basis functions and spectral densities were taken from the EpiNow2 software package (Abbott et al. 2026).

### Forecast evaluation

Forecasting performance was evaluated using the continuous ranked probability score (CRPS) (Gneiting et al. 2007) across the sampled case incidence trajectories produced by each model fit. These scores were calculated across log-transformed case incidence counts (specifically *y* ^*^ log*_e_* (*y* 1)) using the scoringRules R package (Jordan et al. 2019). The log transformation makes score more comparable between different settings and means that forecasts that better predict exponential growth rate (or *R*(*t*)) are rewarded (Bosse et al. 2023).

To calculate skill scores, we produced a baseline model where case incidence follows a random-walk process, with 7-day seasonality imposed to capture any day-of-week effects (i.e. an ARIMA(0, 0, 0)(0, 1, 0)_7_ model). Naive forecasts were produced on the natural scale with a lower truncation applied at zero to prevent negative counts. The magnitude of the random walk was informed by case incidence observations for each location-pathogen pair since January 1 2024, and constraints were applied so that projected incidence was non-negative. Overall skill scores were then calculated as one minus the average CRPS for our forecasts divided by the average CRPS of the baseline forecasts across a given range *X* (defining our aggregation of forecast round, jurisdiction, pathogen, horizon):

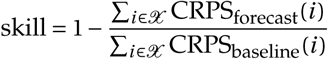

For the sensitivity analyses, forecasts without certain components (e.g. the seasonal Gaussian process) were produced by constraining the value of those components to be zero.

Analysis was performed in R (v4.5.0, R Core 2013) using the tidyverse, patchwork, tidybayes, ggtext, arrow and fabletools packages (Wickham et al. 2019; Pedersen 2025; Kay 2024; Wilke and Wiernik 2022; Richardson et al. 2026; O’Hara-Wild et al. 2026).

## Data and Code Availability

The code used to produce forecasts and figures for this work is available on GitHub at https://github.com/ruarai/reff_comp_public and has been archived at https://doi.org/10.26188/34999054. Anonymised summary data is included to allow for replication of the figures presented. The underlying case datasets used in this work are not publicly available; for access to the raw data, a request must be submitted to the relevant data custodians, which will be assessed independent of the authorship group.

## Acknowledgements

National Notifiable Disease Surveillance System (NNDSS) data were provided by the Interim Australian Centre for Disease Control, Australian Government Department of Health and Aged Care, on behalf of the Communicable Diseases Network Australia (CDNA). We thank public health staff and data managers in state and territory health departments, and the Australian Government Department of Health and Aged Care, along with state and territory public health laboratories.

Public Health Event Surveillance System (PHESS) data were provided by the Department of Health, Victoria. We further wish to acknowledge the medical practitioners and pathology services who diagnosed and notified the cases to the Department of Health, Victoria. We also acknowledge the public health officers, medical officers, epidemiologists and systems specialists who have also contributed to the collection and curation of the PHESS dataset.

The authors would like to thank the members of the ACEFA Hubs Infrastructure Team who processed the case data used in the study. The authors would also like to thank other members of the ACEFA consortium who contributed to valuable discussions during the development of the model.

This research was supported by The University of Melbourne’s Research Computing Services and the Petascale Campus Initiative.

## Supplementary Materials

**Supplementary Table 1:** Estimated multiplicative effects of the between-term and summer school holidays on transmission (*R*(*t*)) and the relative rate of reporting across all five regions and three pathogens considered in the study. Multiplicative effects are calculated as the exponent of the estimated parameter (e.g. exp(*ω*summer)). Values displayed are median and 95% credible interval, with bold values indicating a 95% credible interval that does not overlap one (prior to rounding to two decimal places).

|  |  | Between-term holidays |  | Summer holidays |  |
|---|---|---|---|---|---|
| State/Terr. | Pathogen | *R*(*t*) effect | Reporting effect | *R*(*t*) effect | Reporting effect |
| ACT | RSV | 0.88 (0.75–1.02) | 1.03 (0.91–1.16) | 0.98 (0.75–1.24) | 1.15 (0.81–1.62) |
|  | SARS-CoV-2 | 0.95 (0.86–1.07) | 0.92 (0.80–1.07) | 0.98 (0.86–1.12) | 0.78 (0.58–1.04) |
|  | Influenza | 0.91 (0.82–1.01) | 1.04 (0.90–1.21) | 0.93 (0.79–1.10) | 0.89 (0.63–1.25) |
| NSW | RSV | 0.90 (0.83–0.98) | 0.98 (0.91–1.04) | 0.86 (0.76–0.98) | 0.88 (0.77–1.01) |
|  | SARS-CoV-2 | 0.95 (0.91–0.99) | 0.92 (0.87–0.99) | 0.94 (0.87–1.02) | 1.13 (0.98–1.30) |
|  | Influenza | 0.88 (0.83–0.92) | 0.92 (0.85–0.99) | 0.98 (0.90–1.07) | 0.99 (0.86–1.14) |
| NT | RSV | 1.01 (0.83–1.21) | 1.00 (0.82–1.22) | 0.90 (0.73–1.12) | 0.80 (0.58–1.12) |
|  | SARS-CoV-2 | 0.93 (0.85–1.01) | 0.96 (0.83–1.12) | 0.93 (0.81–1.04) | 0.94 (0.72–1.24) |
|  | Influenza | 0.99 (0.88–1.13) | 0.98 (0.78–1.24) | 0.98 (0.84–1.13) | 0.81 (0.53–1.24) |
| QLD | RSV | 0.92 (0.83–1.01) | 1.00 (0.91–1.08) | 0.92 (0.77–1.08) | 0.96 (0.83–1.10) |
|  | SARS-CoV-2 | 0.97 (0.93–1.02) | 0.99 (0.94–1.05) | 0.92 (0.86–0.99) | 1.05 (0.93–1.18) |
|  | Influenza | 0.93 (0.88–0.99) | 1.01 (0.92–1.09) | 0.89 (0.79–0.99) | 0.96 (0.81–1.15) |
| VIC | RSV | 0.90 (0.83–0.97) | 0.96 (0.88–1.03) | 0.87 (0.76–0.99) | 0.76 (0.64–0.89) |
|  | SARS-CoV-2 | 0.95 (0.90–1.00) | 0.95 (0.87–1.01) | 0.91 (0.82–1.01) | 0.84 (0.73–0.97) |
|  | Influenza | 0.87 (0.82–0.92) | 0.98 (0.88–1.08) | 0.93 (0.86–1.02) | 0.73 (0.62–0.86) |

**Supplementary Figure 1:**
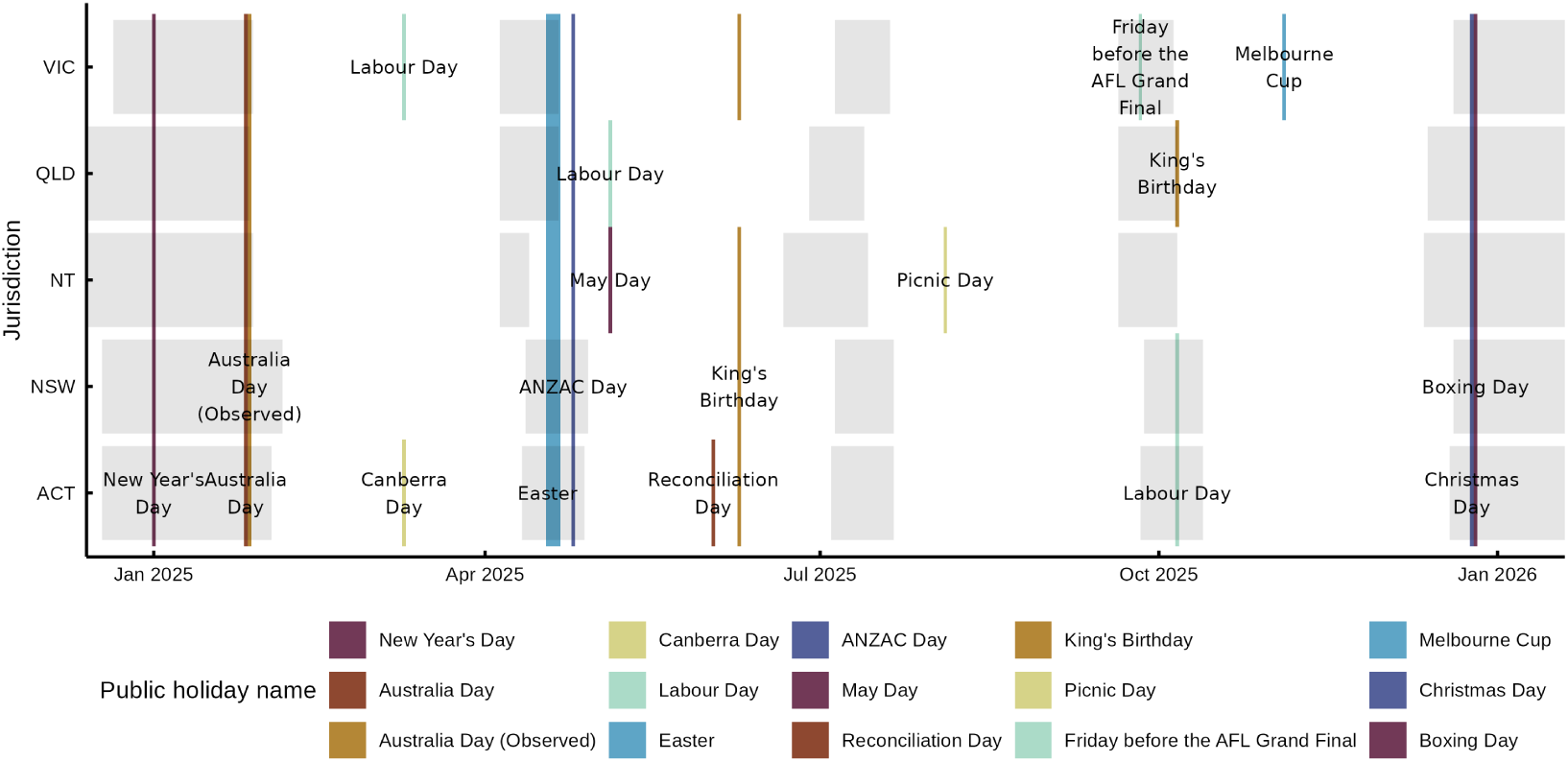
Public and school holidays across the five jurisdictions of Australia considered in the study. School holidays are indicated in light grey shading.

**Supplementary Figure 2:**
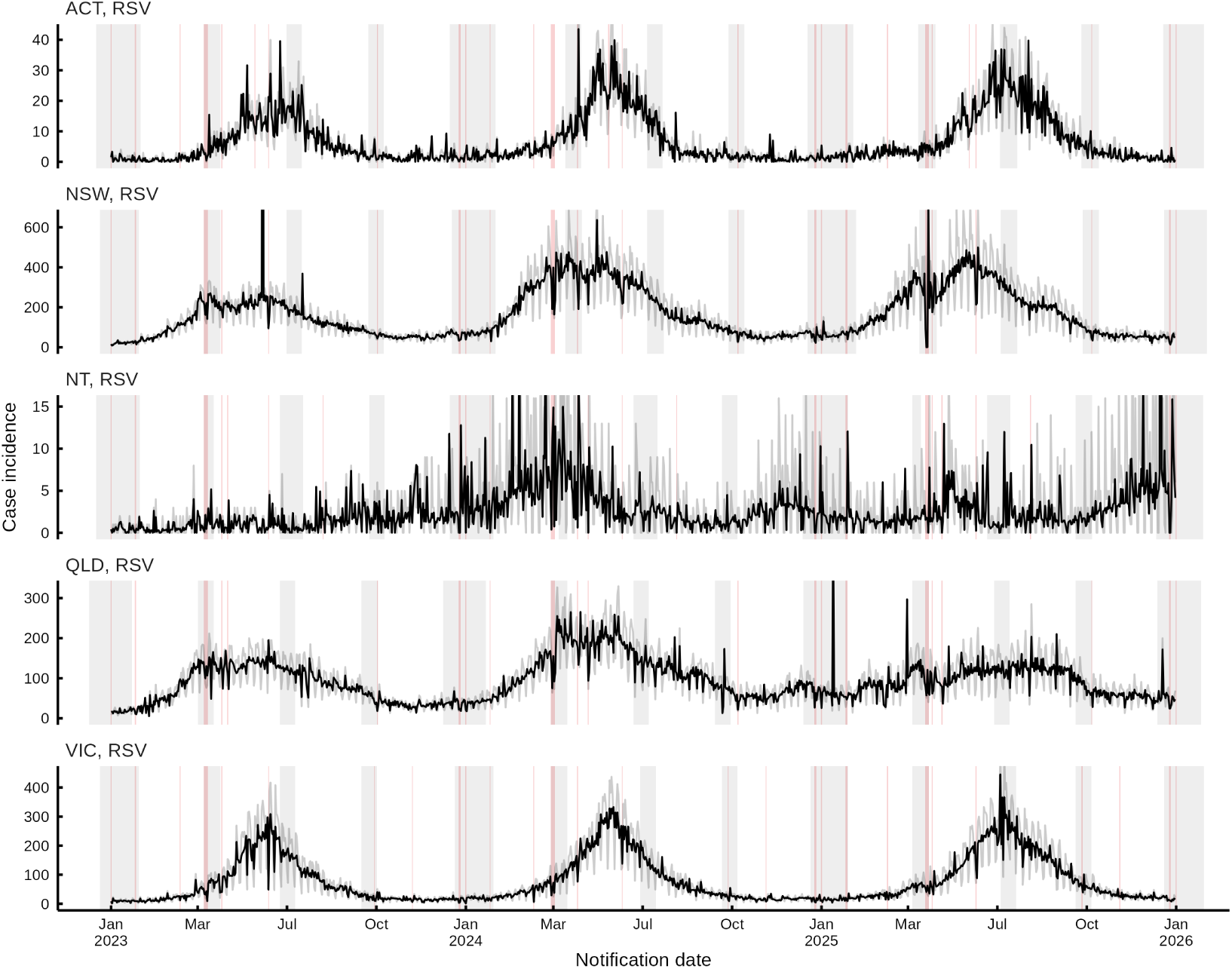
Daily case incidence (light grey) and STL smoothed case incidence (black) for respiratory syncytial virus (RSV) for the five jurisdictions considered in the study. Data are displayed back to the start of the fitting period for RSV. School holidays are indicated in light grey shading while public holidays are indicated in light red.

**Supplementary Figure 3:**
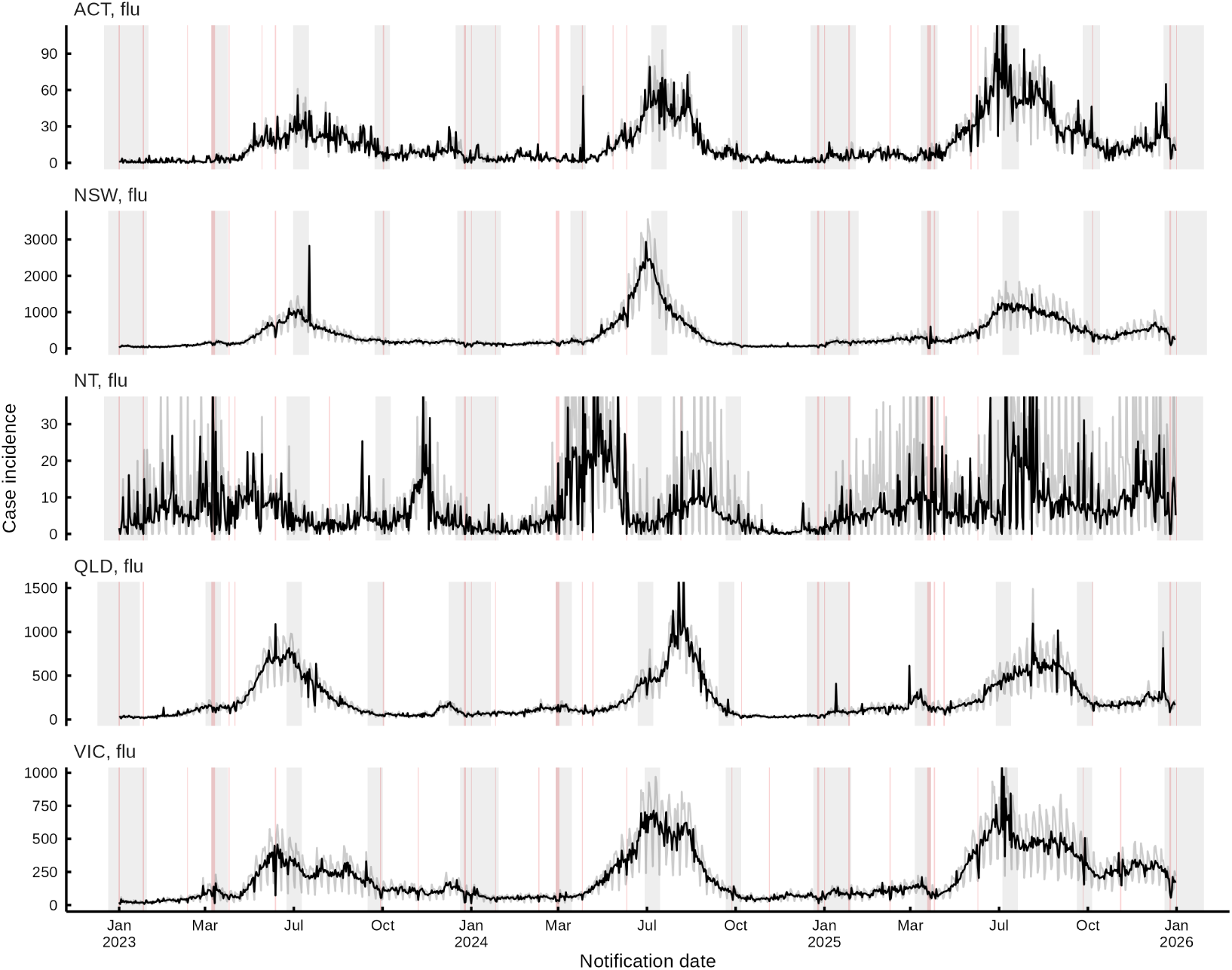
Daily case incidence (light grey) and STL smoothed case incidence (black) for influenza for the five jurisdictions considered in the study. Data are displayed back to the start of the fitting period for influenza. School holidays are indicated in light grey shading while public holidays are indicated in light red.

**Supplementary Figure 4:**
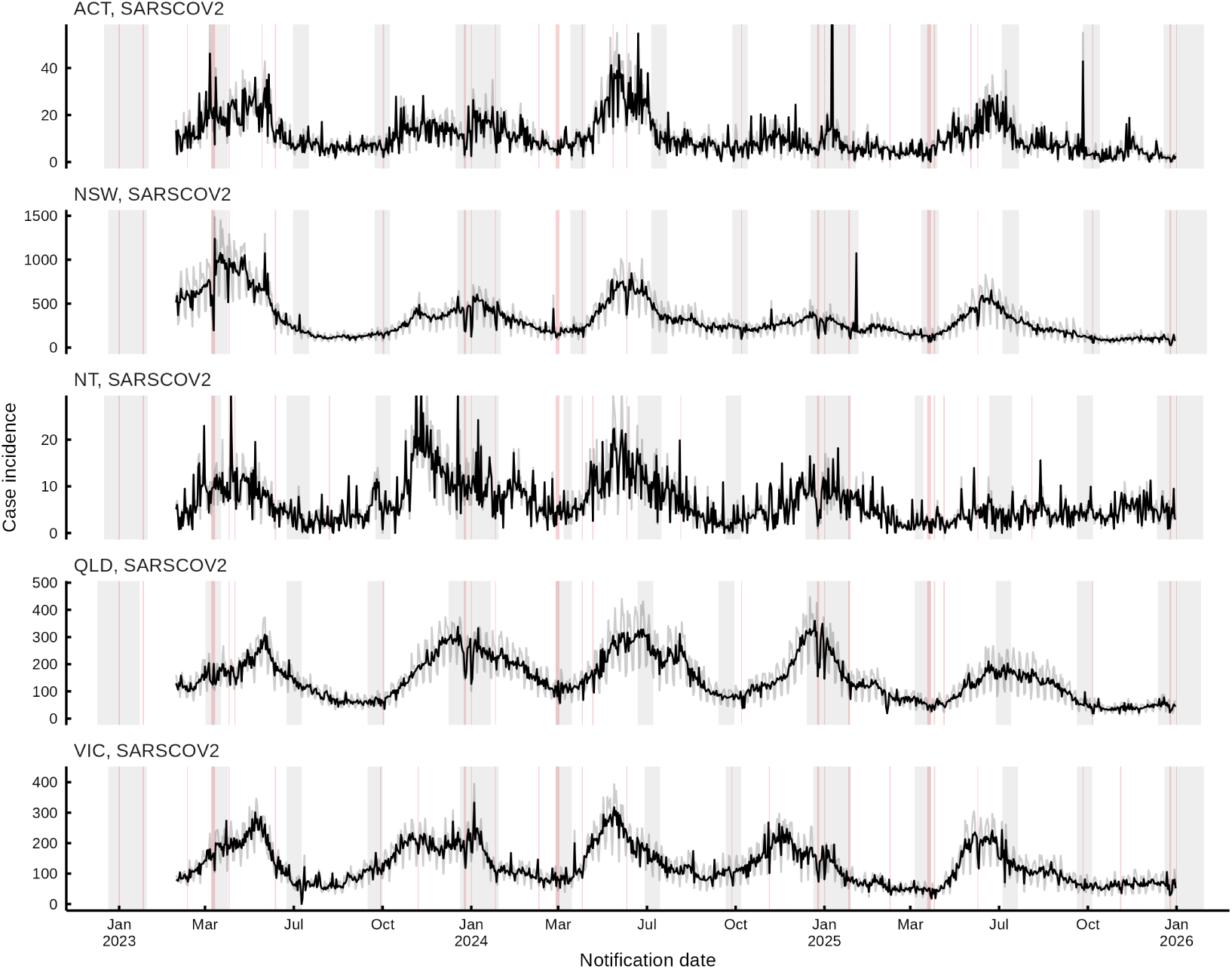
Daily case incidence (light grey) and STL smoothed case incidence (black) for SARS-CoV-2 for the five jurisdictions considered in the study. Data are displayed back to the start of the fitting period for SARS-CoV-2. School holidays are indicated in light grey shading while public holidays are indicated in light red.

**Supplementary Figure 5:**
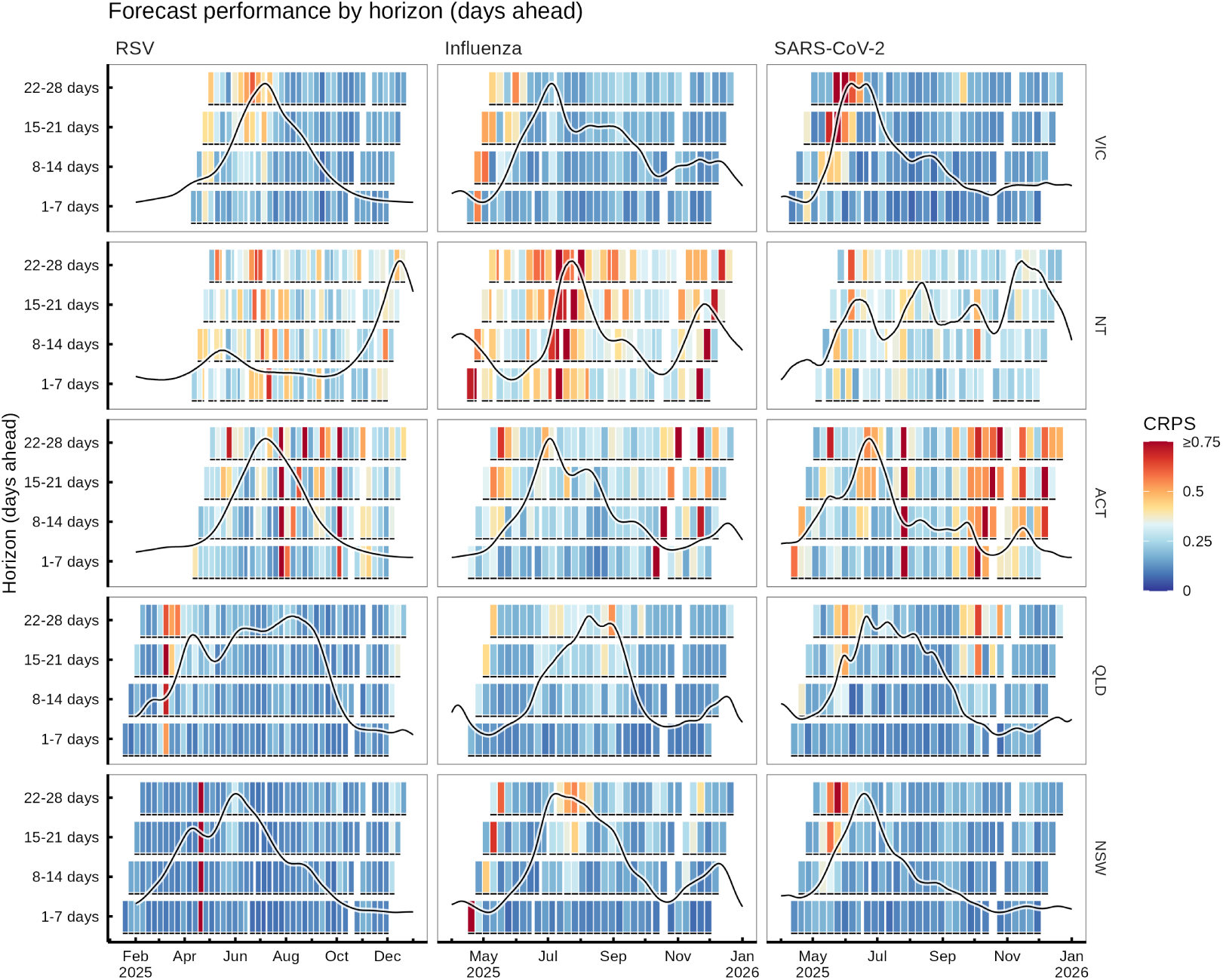
Extension of Figure 2 to examine performance at different forecast horizons from one-week ahead to four-weeks ahead. As in Figure 2, STL smoothed case incidence is plotted over the CRPS (with scores aligned to the corresponding date of the nth week of the forecast) to allow for comparison of performance to the underlying epidemiological activity. Each forecast forms a diagonal line of squares: for example, the last influenza forecast for NT was uniformly poor across the four weeks. In contrast, a vertical line of poor performance (as in e.g. NSW RSV) indicates that four consecutively produced forecasts failed to predict that week of data well.

**Supplementary Figure 6:**
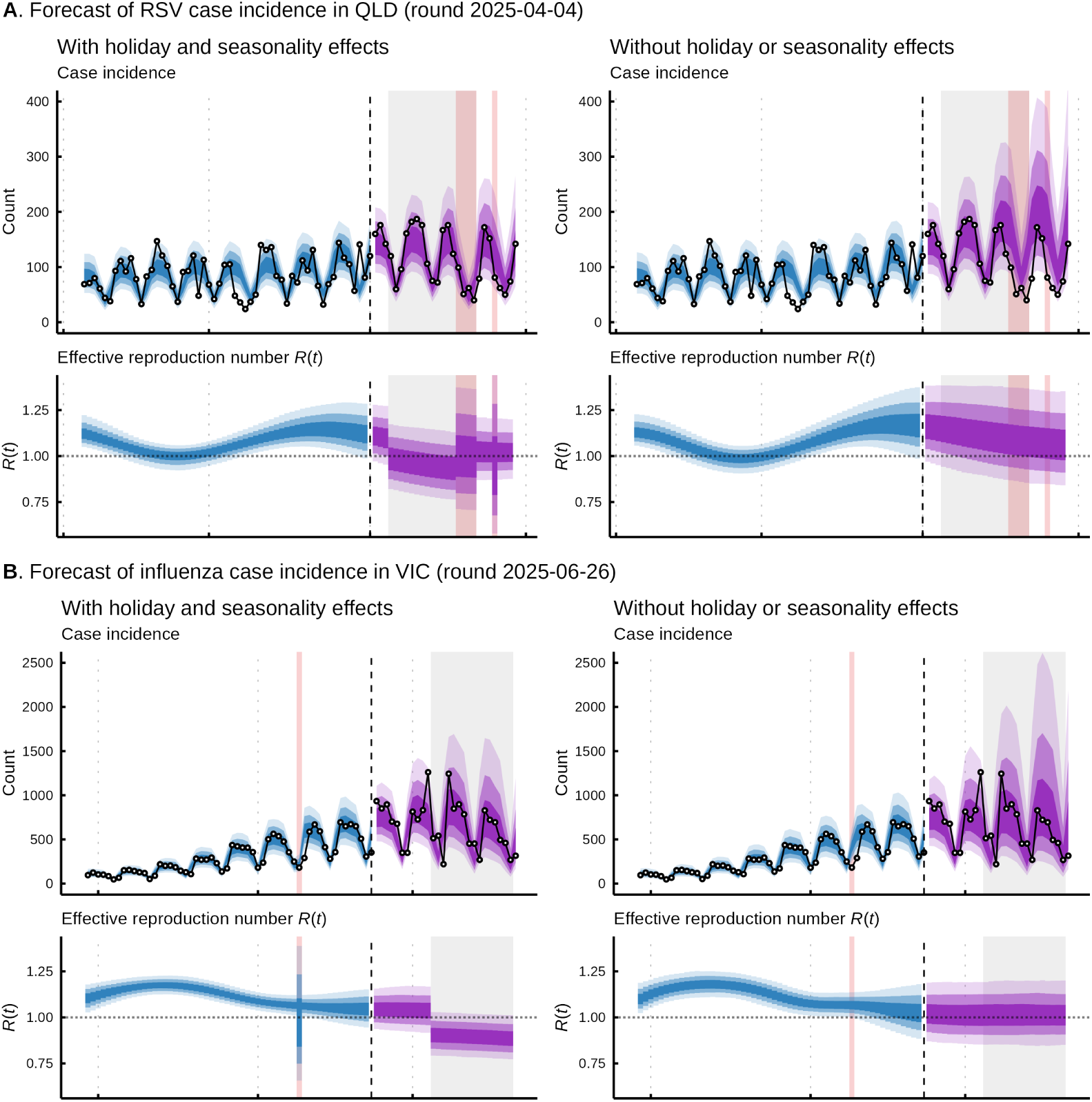
A comparison of forecasts produced with (left) and without (right) the holiday (including school and public holiday effects across both transmission and the relative rate of reporting) and seasonality transmission effects. 50%, 80%, and 95% intervals are displayed. **A**. RSV forecasts in QLD for round 2025-04-04. **B**. Influenza forecasts in VIC for round 2025-06-26.

**Supplementary Figure 7:**
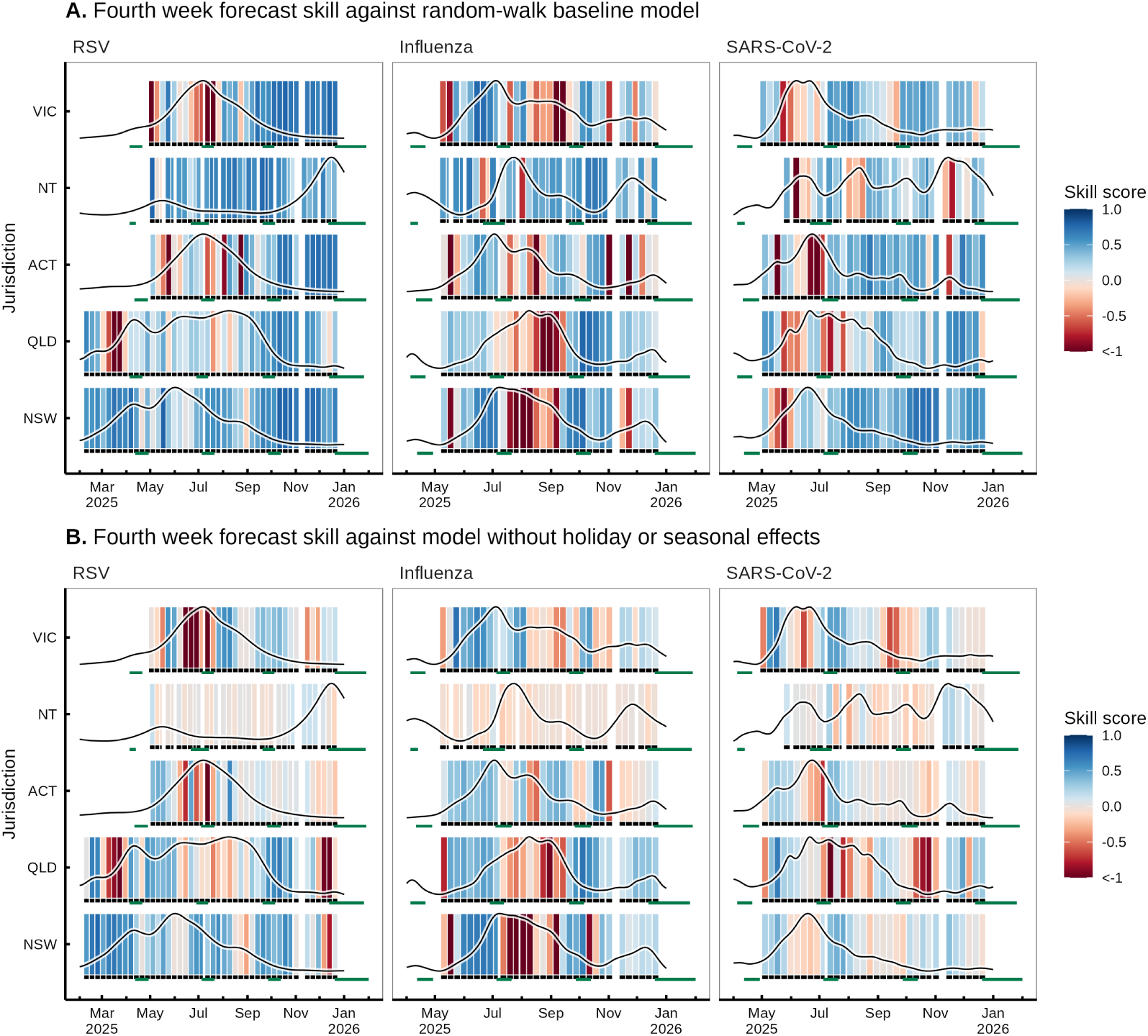
Extension of Figure 2 to examine forecast skill in comparison to the randomwalk baseline (**A**, see Methods) or the model without holiday and seasonality effects (**B**, as in Figure 4) at the four-week forecast horizon. School holiday periods are underlined in green for each jurisdiction. As in Figure 2, STL smoothed case incidence is plotted over the skill score (with scores aligned to the corresponding date of the fourth week of the forecast) to allow for comparison of performance to the underlying epidemiological activity.

**Supplementary Figure 8:**
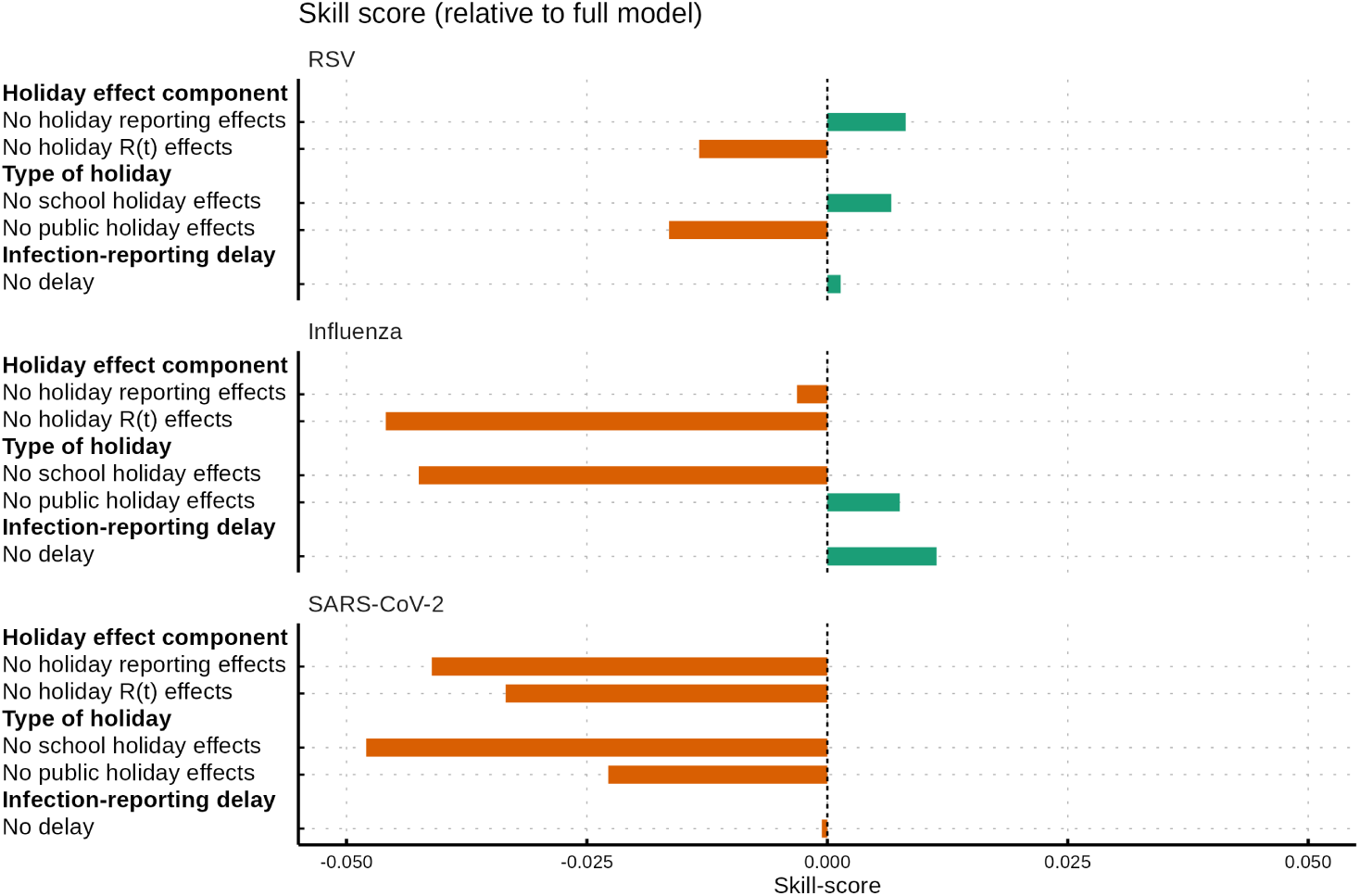
The impact of removing different model components on the average forecasting performance as calculated via skill score. Skill scores were calculated from CRPS across log-counts for the fourth week horizon. Each row indicates the performance of a model with a certain component removed from the model compared to the ‘full’ model (i.e. that described in the main text). A negative skill score indicates that removing that component reduces forecasting performance, whereas a positive skill score indicates that removing the component improves performance.

**Supplementary Figure 9:**
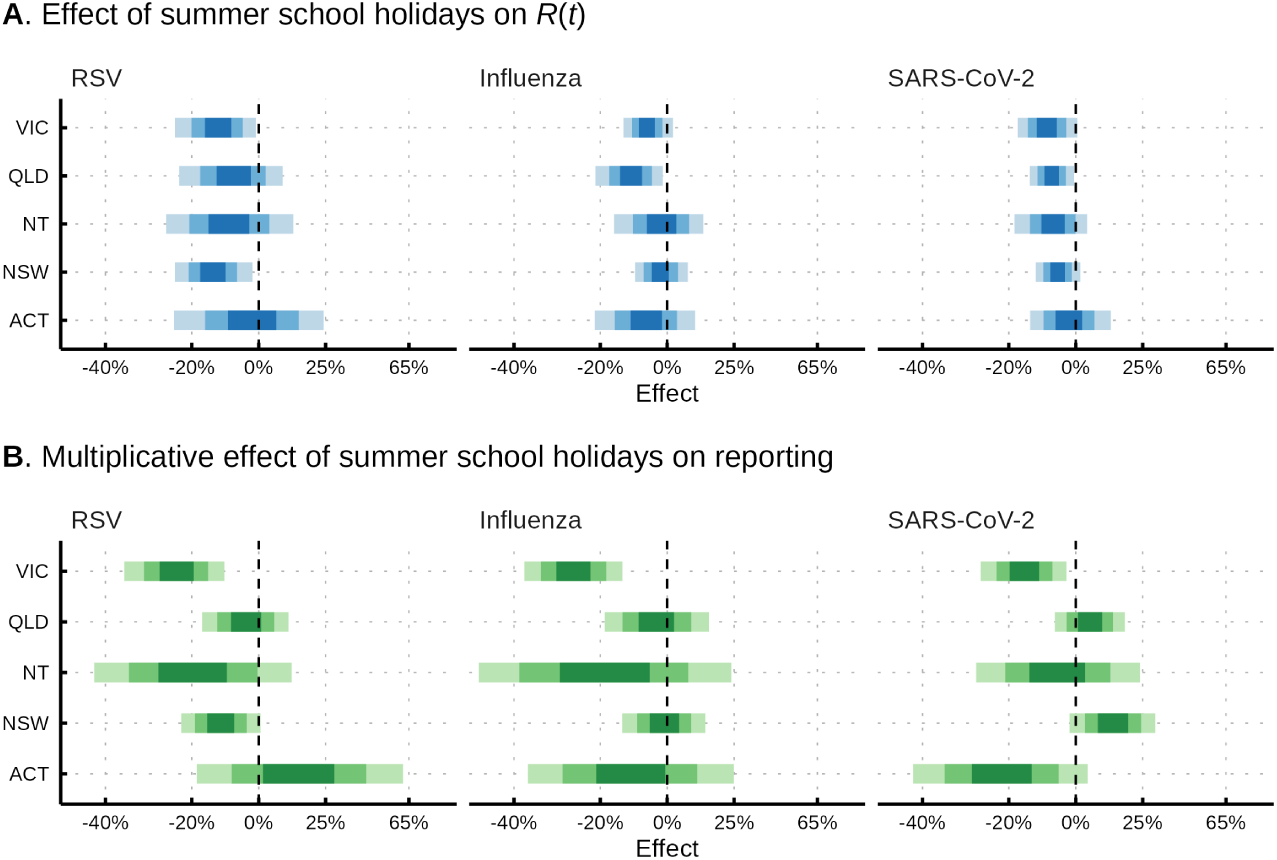
Extension of Figure 5 illustrating the effect of the summer holidays on transmission (*R*(*t*)) (**A**) and the relative rate of reporting (**B**). 50%, 80% and 95% credible intervals are displayed.

**Supplementary Figure 10:**
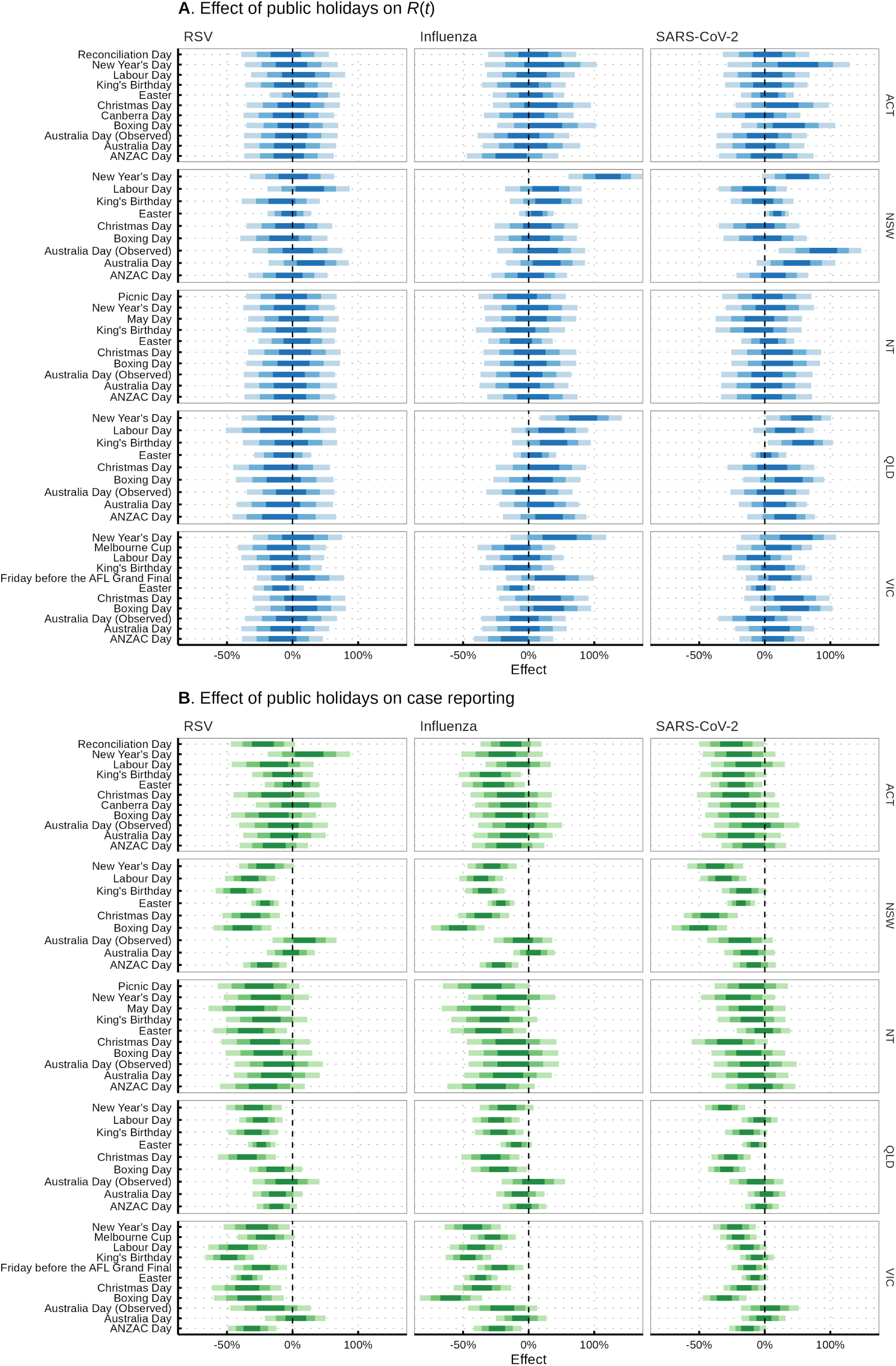
Extension of Figure 5 illustrating the effect of public holidays on transmission (*R*(*t*)) (**A**) and the relative rate of reporting (**B**). 50%, 80% and 95% credible intervals are displayed.

**Supplementary Figure 11:**
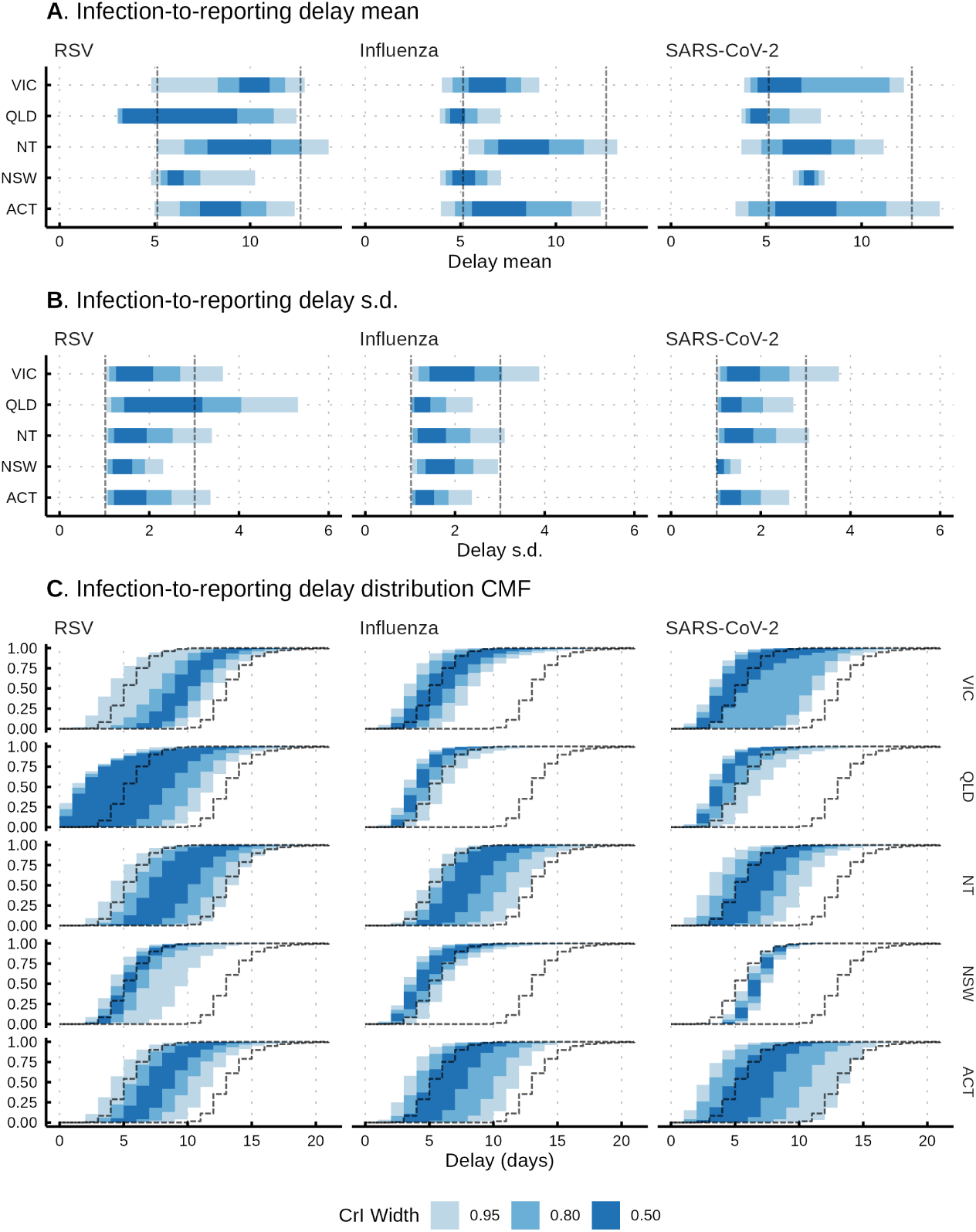
Estimated delay between infection and case reporting. 50%, 80% and 95% credible intervals are displayed. **A**, **B**. Mean and standard deviation of the log-normal delay distribution. **C**. Cumulative mass functions (CMF) of the delay distributions across the posterior with pointwise credible intervals. Dashed lines indicate the 95% credible intervals of the prior for the delay distribution.

**Supplementary Figure 12:**
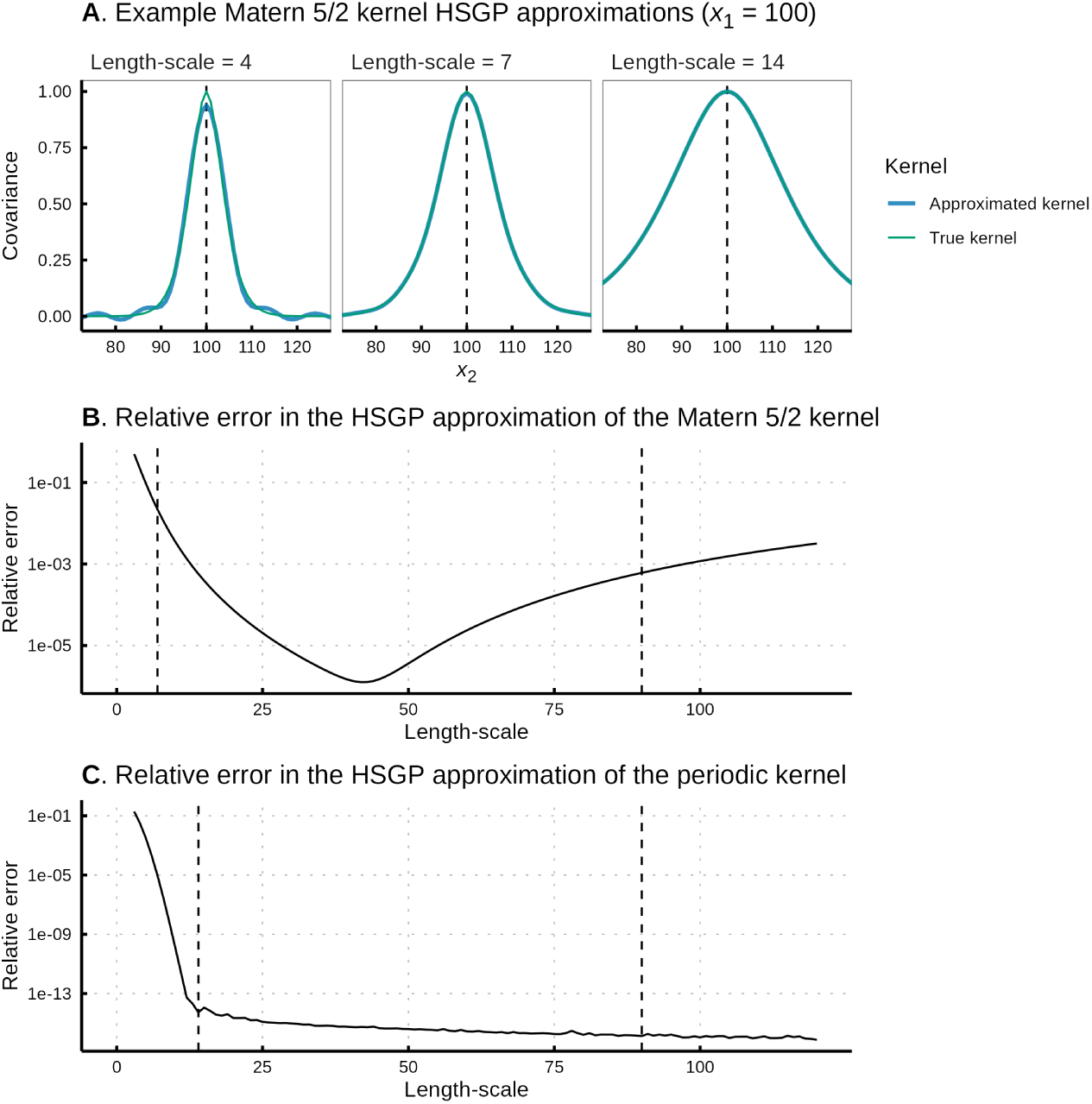
Validation of the HSGP approximation of the Matérn 5/2 and periodic Gaussian processes used in the model. Approximations are produced across 365 3 time-points and with the number of basis functions as described in the main text. **A**. Example Matérn 5/2 kernels for varied length-scale calculated exactly or under the HSGP approximation. **B**. The relative error between the approximated kernel and true kernel (see Equation 18 in Riutort-Mayol et al. 2023). The vertical dashed lines indicate the 99% interval on the prior for length-scale (7 and 90 days). **C**. As before, but for the periodic (seasonal) Gaussian process. The vertical dashed lines indicate the 99% interval on the prior for length-scale (14 and 90 days).

**Supplementary Figure 13:**
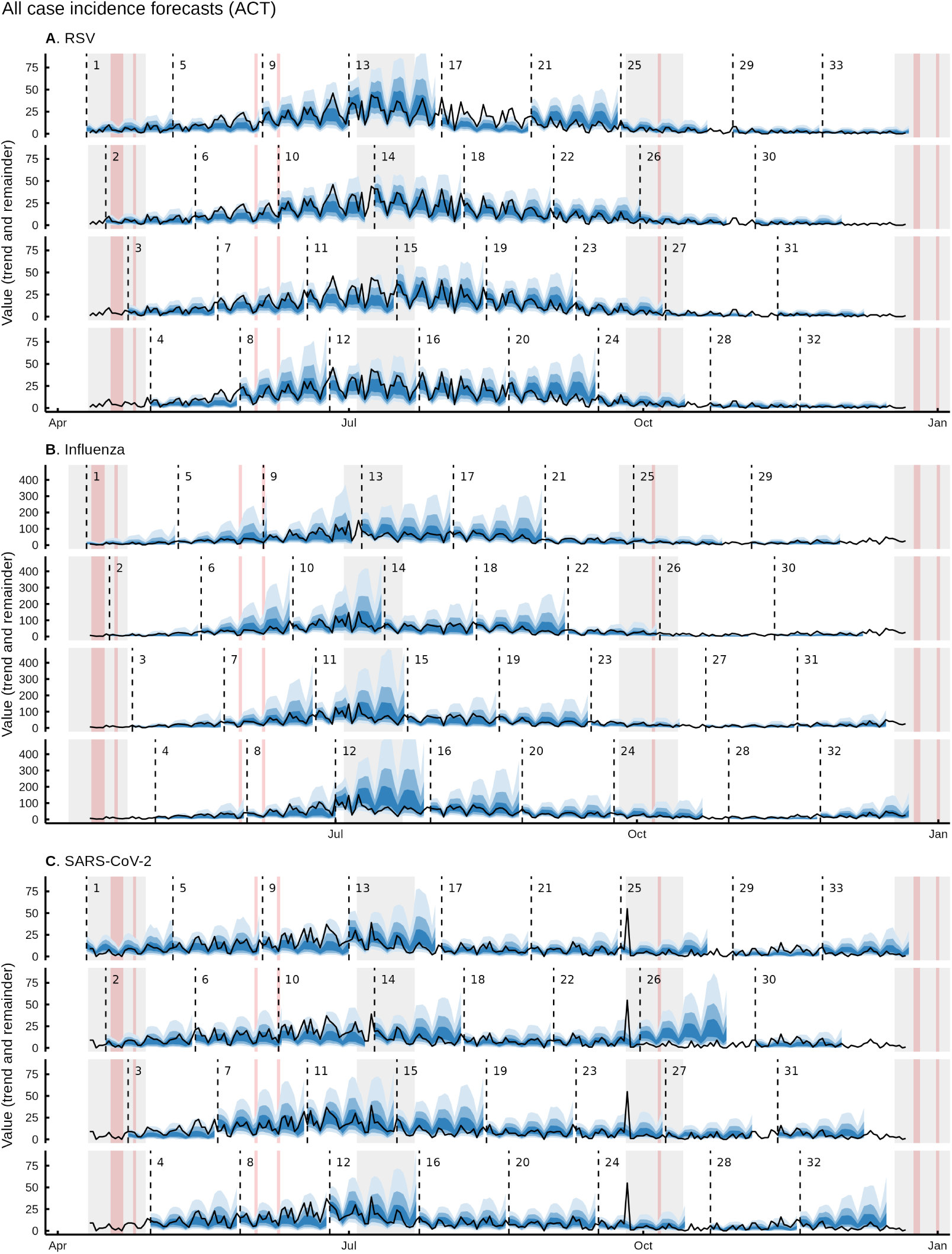
All case incidence forecasts for RSV, influenza and SARS-CoV-2 in the Australian Capital Territory. School holidays are shaded in the background in grey and public holidays in red. 50%, 80% and 95% credible intervals are displayed.

**Supplementary Figure 14:**
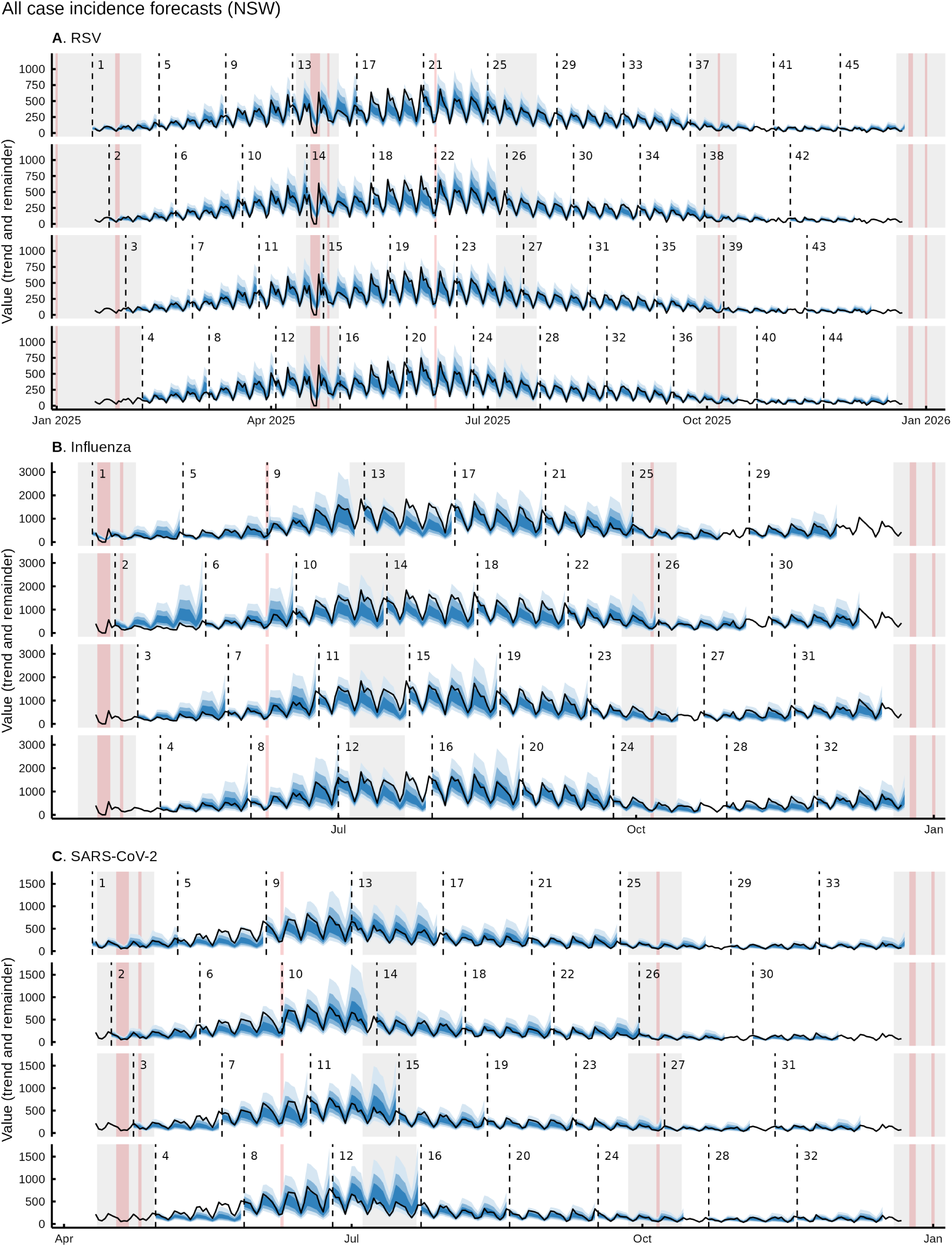
All case incidence forecasts for RSV, influenza and SARS-CoV-2 in New South Wales. School holidays are shaded in the background in grey and public holidays in red. 50%, 80% and 95% credible intervals are displayed.

**Supplementary Figure 15:**
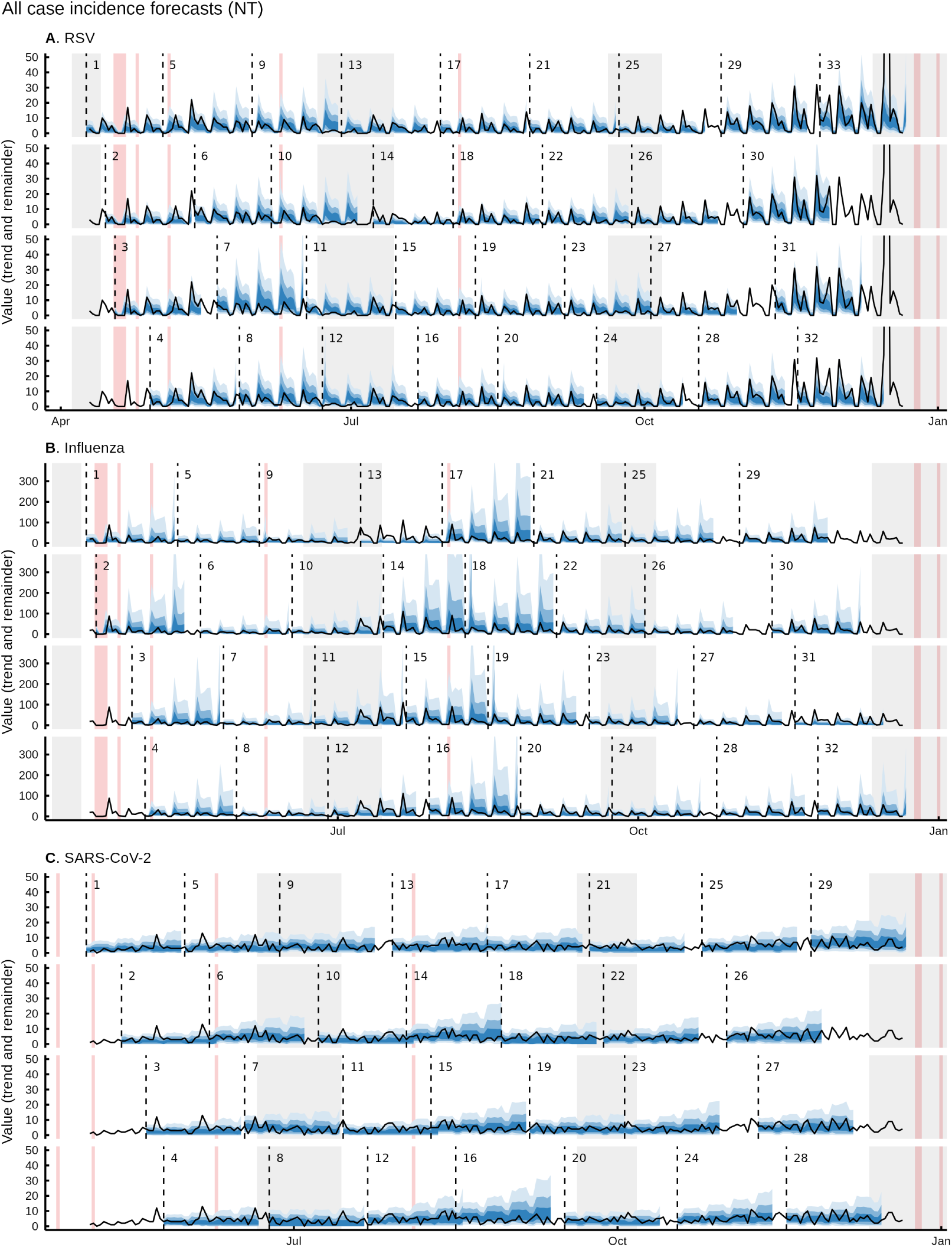
All case incidence forecasts for RSV, influenza and SARS-CoV-2 in the Northern Territory. School holidays are shaded in the background in grey and public holidays in red. 50%, 80% and 95% credible intervals are displayed.

**Supplementary Figure 16:**
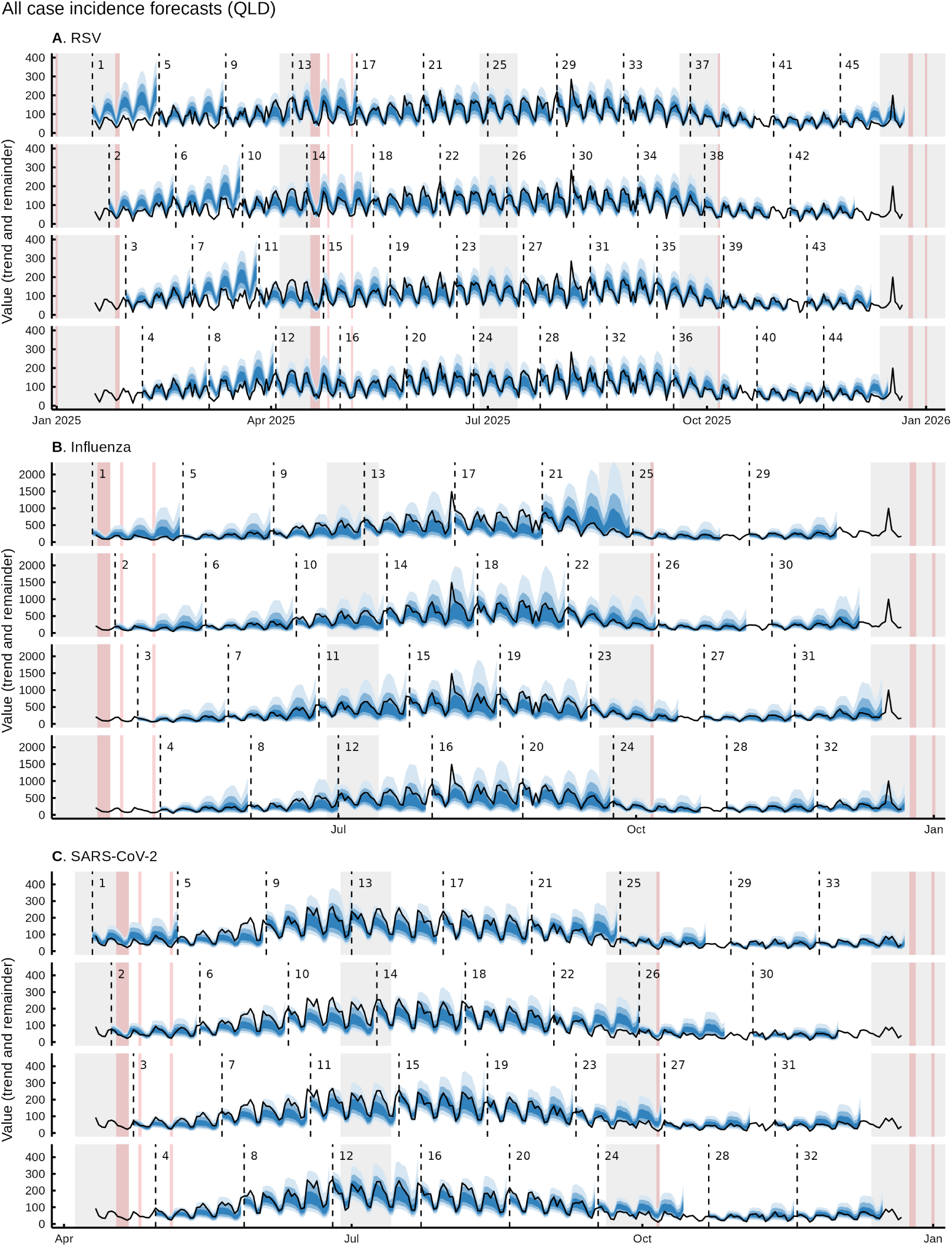
All case incidence forecasts for RSV, influenza and SARS-CoV-2 in Queensland. School holidays are shaded in the background in grey and public holidays in red. 50%, 80% and 95% credible intervals are displayed.

**Supplementary Figure 17:**
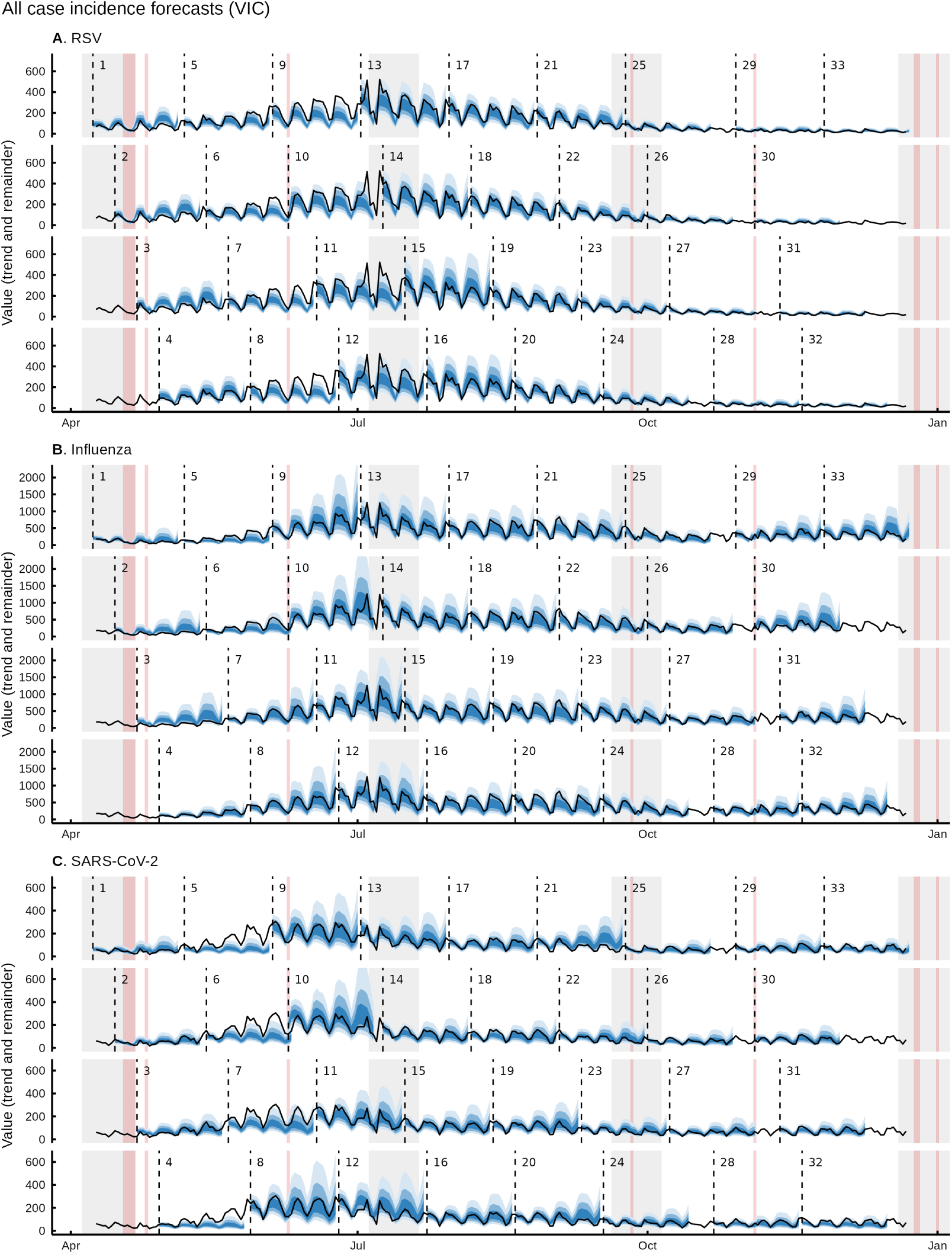
All case incidence forecasts for RSV, influenza and SARS-CoV-2 in Victoria. School holidays are shaded in the background in grey and public holidays in red. 50%, 80% and 95% credible intervals are displayed.

